# Leveraging genomic diversity for discovery in an EHR-linked biobank: the UCLA ATLAS Community Health Initiative

**DOI:** 10.1101/2021.09.22.21263987

**Authors:** Ruth Johnson, Yi Ding, Vidhya Venkateswaran, Arjun Bhattacharya, Alec Chiu, Tommer Schwarz, Malika Freund, Lingyu Zhan, Kathryn S. Burch, Christa Caggiano, Brian Hill, Nadav Rakocz, Brunilda Balliu, Jae Hoon Sul, Noah Zaitlen, Valerie A. Arboleda, Eran Halperin, Sriram Sankararaman, Manish J. Butte, UCLA Precision Health Data Discovery Repository Working Group, UCLA Precision Health ATLAS Working Group, Clara Lajonchere, Daniel H. Geschwind, Bogdan Pasaniuc

## Abstract

Large medical centers located in urban areas such as Los Angeles care for a diverse patient population and offer the potential to study the interplay between genomic ancestry and social determinants of health within a single medical system. Here, we introduce the UCLA ATLAS Community Health Initiative – a biobank of genomic data linked with de-identified electronic health records (EHRs) of UCLA Health patients. We leverage the unique genomic diversity of the patient population in ATLAS to explore the interplay between self-reported race/ethnicity and genetic ancestry within a disease context using phenotypes extracted from the EHR. First, we identify an extensive amount of continental and subcontinental genomic diversity within the ATLAS data that is consistent with the global diversity of Los Angeles; this includes clusters of ATLAS individuals corresponding to individuals with Korean, Japanese, Filipino, and Middle Eastern genomic ancestries. Most importantly, we find that common diseases and traits stratify across genomic ancestry clusters, thus suggesting their utility in understanding disease biology across diverse individuals. Next, we showcase the power of genetic data linked with EHR to perform ancestry-specific genome and phenome-wide scans to identify genetic factors for a variety of EHR-derived phenotypes (phecodes). For example, we find ancestry-specific associations for liver disease, and link the genetic variants with neurological and neoplastic phenotypes primarily within individuals of admixed ancestries. Overall, our results underscore the utility of studying the genomes of diverse individuals through biobank-scale genotyping efforts linked with EHR-based phenotyping.

## Introduction

Linking electronic health records (EHRs) to patient genomic data within biobanks in a de-identified fashion has the potential to significantly advance genomic discoveries and precision medicine efforts (e.g., population screening, identifying drug targets)[1]–[4]. However, the underrepresentation of minoritized populations in biomedical research [5]–[11]raises concerns that advancements in precision medicine may widen disparities in access to high-quality health care [12]–[14]. For example, European-ancestry individuals constitute approximately 16% of the global population, yet account for almost 80% of all genome-wide association study (GWAS) participants [13]. As a direct result of this imbalance, existing methods to predict disease risk from genetics (e.g., polygenic risk scores) are vastly inaccurate in individuals of non-European ancestry [13], [15] thus forming a barrier for advancing genomic medicine to benefit patients of all ancestries.

The UCLA Health medical system is located in Los Angeles, one of the most ethnically diverse cities in the world. There is no ethnic majority: 48.5% of Los Angeles residents self-identify as Hispanic or Latino, 11.6% as Asian, and 8.9% as Black or African American; additionally, 37% of Los Angeles residents are neither U.S. nationals, nor U.S. citizens at birth [16]. Therefore, the UCLA Health patient population and the availability of digital health data captured in EHRs from a single medical system present a unique opportunity to increase the inclusion of underrepresented minorities in biomedical research. We introduce the UCLA ATLAS Community Health Initiative (or ATLAS for brevity), a biobank embedded within the UCLA Health medical system composed of de-identified, EHR-linked genomic data from a diverse patient population. The current initiative aims to collect data from over 150,000 individuals; currently this consists of 26,414 individuals genotyped at 673,148 variants genome-wide each using the Illumina global screening array (GSA) [17]. The EHR contains a de-identified extract of medical records (billing codes, laboratory values, etc.) as well as demographic information such as self-reported race and ethnicity information. It is important to note that self-reported race and ethnicity (SIRE) represent social constructs that capture shared values, cultural norms, and behaviors of subgroups [18] that are distinct concepts from genetic ancestry, which refers to the history of one’s genome with little to no relation to cultural aspects of identify. This difference is even more relevant for individuals self-describing as multi-racial (and/or admixed) where genetic ancestry bears little correlation to SIRE [19], [20]. Understanding the interplay of genetic factors (such as genetic ancestry) with social determinants of health (as inferred from self-reports) is still mired in the confounding overlaps between race, socioeconomic status, and disease, but serves as a critical step in mapping and predicting disease risk across individuals of all ancestries, thus enabling equitable genomic medicine to individuals of all ancestries.

In this work, we leverage the unique genomic diversity of the patient population in ATLAS to explore the interplay between self-reported race/ethnicity and genetic ancestry within a disease context using phenotypes extracted from the EHR within a single medical system. We cluster individuals by genetic ancestry within the EHR-linked biobank, systematically construct phenotypes from EHR, and compute disease associations using multi-ethnic pipelines for both genome-wide and phenome-wide association studies. We find that genetic ancestry and self-reported ancestry yield distinct subpopulations thus emphasizing the distinction between genetic ancestry and self-reported race and ethnicities. We leverage genetic and self-reported data to find extensive variation of sub-continental ancestry within ATLAS across European, Asian, and American ancestries. For example, we find clusters of individuals with recent ancestry from Filipino, Japanese, and Korean ancestries. Such sub-continental clusters also stratify individuals according to disease groups thus emphasizing their utility in biomedical research. We perform genome-wide and phenome-wide association studies to recapitulate known genomic risk regions; as an example, focusing on chronic nonalcoholic liver disease, we recapitulate the 22q13.31 locus and perform a phenome-wide association study across 1,330 EHR-derived phenotypes at the lead SNP, rs2294915, across multiple populations. We describe genetic associations for liver-related phenotypes in multiple ancestry groups as well as associations with neurological and neoplastic phenotypes that are associated exclusively in the Admixed American group. These results underscore how the utility of large-scale genetic analyses and deep phenotyping in diverse populations have substantial medical relevance for population health.

## Results

### ATLAS includes individuals of diverse continental ancestries

The UCLA Health patient population is diverse, with 63.3% self-reporting their race as White or Caucasian, 6.7% as Black or African American, 10.5% as Asian, 0.6% as American Indian or Alaska Native, 0.3% as Pacific Islander, and 18.6% identify as one of the additional races listed in detail in the Supplementary Materials (Figure 1A, Supplementary Table S1, S3). 15.8% of individuals self-report their ethnicity as Hispanic or Latino; the remaining individuals self-report as non-Hispanic/Latino (Figure 1A, Supplementary Table S2, S3). We investigated genetically inferred ancestry through principal component analysis (PCA)[21], [22], to identify population clusters according to the five continental “superpopulations” defined in the 1000 Genomes reference panel [23] (see Methods; Figure 1B, 2A; Supplementary Figure S1, S2; Supplementary Table S4). Although we broadly find that SIRE is concordant with the inferred continental genetic ancestry, we find marked differences between genetically defined ancestry groups and SIRE, further emphasizing that genetic ancestry is a distinct concept from self-reported race and ethnicity. For example, we find >10% of individuals within the European genetic ancestry group do not identify as Non-Hispanic/Latino – White/Caucasian (NH-WC) SIRE; 10% of individuals within the African genetic ancestry group do not self-report as Non-Hispanic/Latino – Black/African American (NH-AfAm), and >25% of the Admixed American genetic ancestry group do not identify as Hispanic/Latino – Other Race (HL-Other) or Hispanic/Latino – White/Caucasian (HL-WC) (Supplementary Table S5).

**Figure 1:**
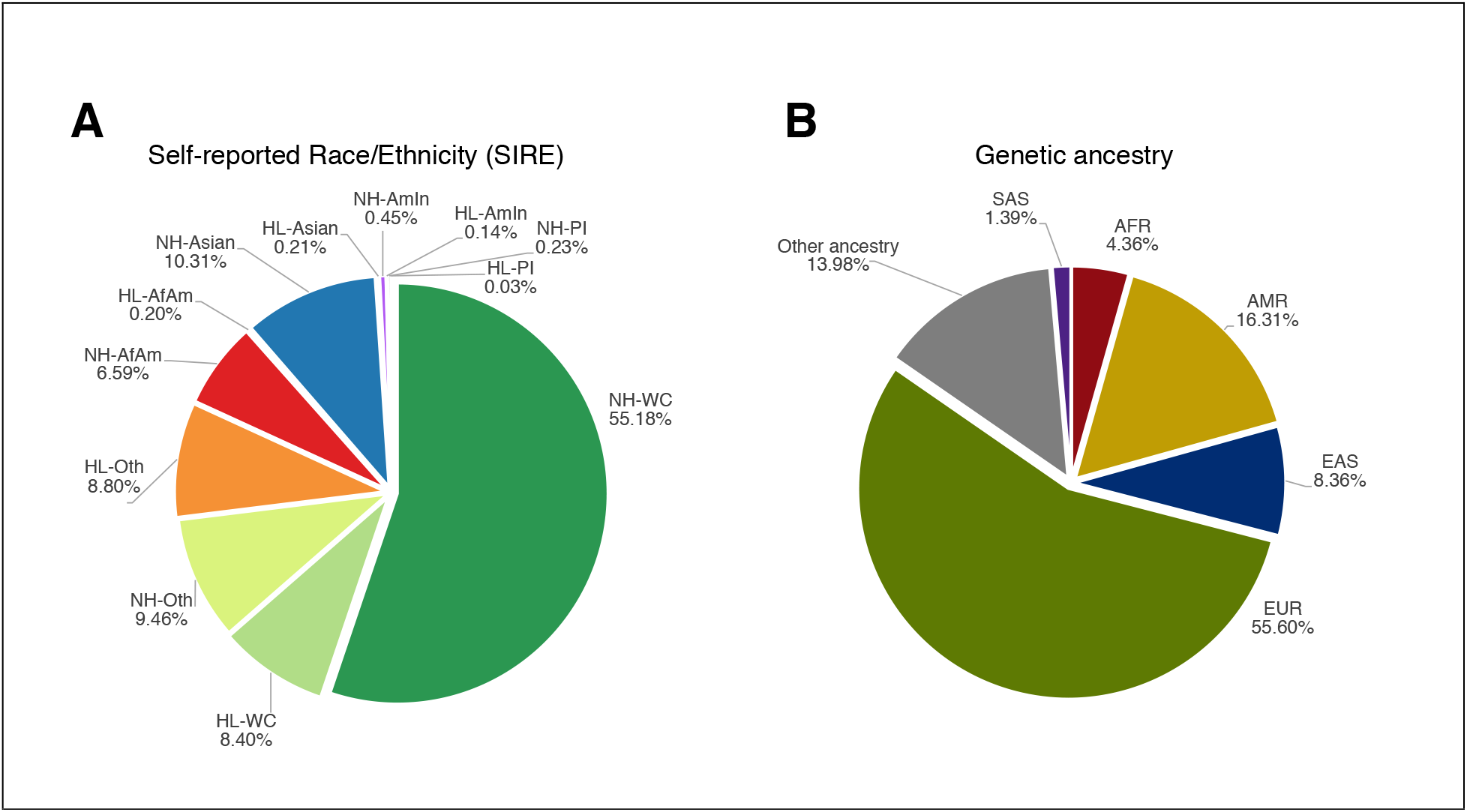
Self-reported race/ethnicity (SIRE) and genetic ancestry capture distinct information. We show the percentage breakdown of (A) all SIREs and (B) continental genetic ancestry for all unrelated individuals in ATLAS (N= 25,842). We exclude individuals whose self-reported race and/or ethnicity are unknown.

Further making the distinction between genetic ancestry and SIRE, we reveal extensive genetic heterogeneity both between and within SIREs within orthogonal spectra from PCA (Figure 2A and 2B). For example, most individuals who self-report as NH-AfAm lie along a cline between the African and European genetic ancestry clusters. We also observe that the cluster of individuals with inferred African ancestry from PCA form a considerably smaller cluster than the group of individuals in the NH-AfAm SIRE in ATLAS. Within ATLAS, we find that 1,426 individuals self-identify as NH-AfAm, but only 1,233 individuals are grouped into the African genetic ancestry cluster. This difference is likely because many individuals in ATLAS identify as African American, which suggests genetic admixture between African and European ancestry in this group. Conversely, there are fewer individuals in the Non-Hispanic/Latino – Asian (NH-Asian) SIRE (N=2,469) than those grouped into the East Asian and South Asian ancestry clusters (N= 2,611). A similar trend follows for the NH-WC SIRE (N= 14,328) and the European ancestry cluster (N= 14,800). The majority of individuals who are included in the genetic ancestry clusters, but not the corresponding SIREs, had either unknown SIRE information or reported their race as ‘Other Race’, demonstrating how genetic ancestry inference can be advantageous when self-reported information is not known or individuals’ race/ethnicity are not represented in patient questionnaires. However, 14% of individuals still have unclassifiable genetic ancestry (Supplementary Table S4) either because they are clustered into multiple ancestry groups or no ancestry group as all. The latter could be due to extensive admixture in their genomes or the absence of relevant ancestral groups in the chosen reference panels.

**Figure 2:**
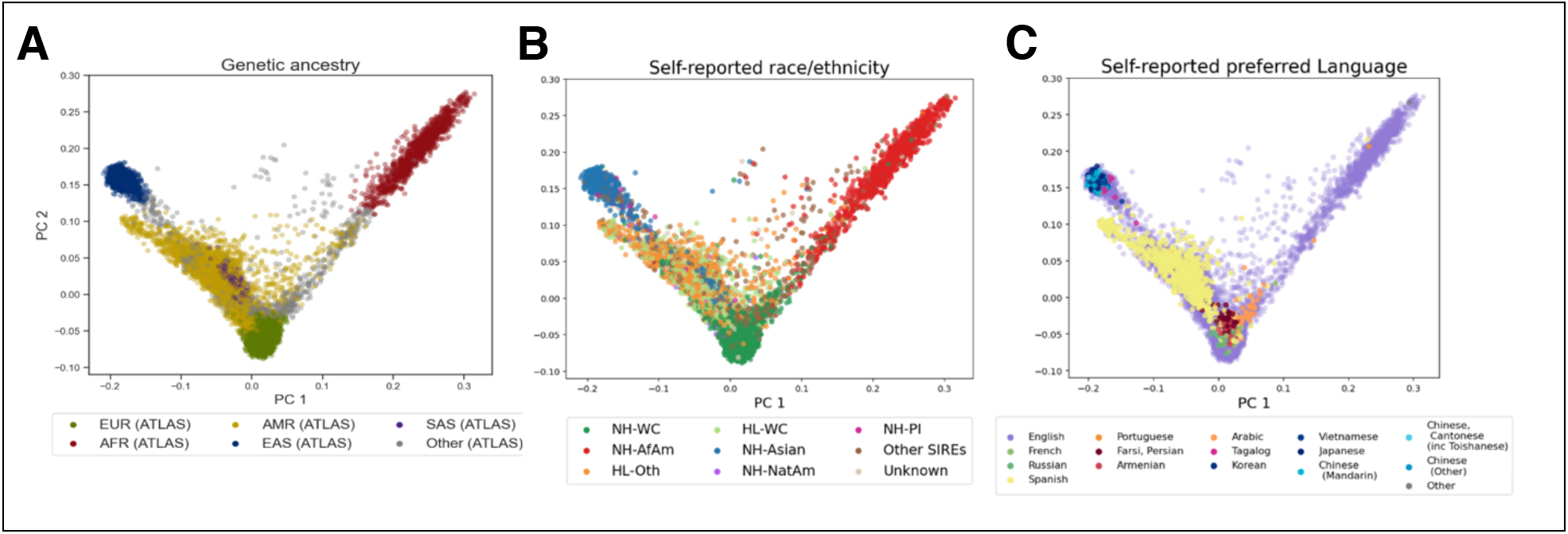
Global PCA reflects self-reported race/ethnicity and preferred language of ATLAS participants. (A) Genetic PCs 1 and 2 of individuals in ATLAS (N=25,842) shaded by inferred continental genetic ancestry (European, African, Admixed American, East Asian, and South Asian) as inferred from 1000 Genomes. (B) and (C) show the first two genetic PCs of the ATLAS participants shaded by SIRE and preferred language, respectively. To improve visualization in (C), only languages with >10 responses are assigned a color.

Labeling individuals by self-reported preferred language, we observe trends that are consistent with both SIRE and continental genetic ancestry (Figure 2C). For example, out of all individuals who report Spanish as their primary language, 96.1% of these individuals were estimated to have Admixed American genetic ancestry. Additionally, 98.5% of individuals who report Japanese, Korean, Tagalog, Vietnamese, Mandarin Chinese, and Cantonese as their primary languages were inferred to have East Asian genetic ancestry. We also see clusters of individuals who speak Armenian, Arabic, and Farsi/Persian; we find that 26.2% of the individuals that speak these languages could not be classified into one of the five continental ancestry groups within 1000 Genomes. This discrepancy is likely because the 1000 Genomes reference panel does not contain samples from regions where these languages are primarily spoken. These findings showcase the limitation of current reference panels of genetic diversity and demonstrate the value of characterizing individuals using both genetic ancestry and self-reported information.

### Fine-scale subcontinental ancestry within ATLAS individuals

Next, we inferred the ancestry of individuals within the ATLAS East Asian ancestry group (EAS) and identified 5 subcontinental genetic ancestry groups (Figure 3A, Supplementary Figure S3) [23]. First, we clustered individuals in ATLAS according to 3 different subgroups of Chinese ancestry (Han Chinese, Southern Han Chinese, and Dai Chinese). Additionally, when projecting individuals’ preferred language onto the PCs, two distinct clusters are delineated according to Chinese Mandarin and Chinese Cantonese/Toishanese (Supplementary Figure 5B). The Southern Han Chinese cluster of individuals highly correlates with individuals speaking Chinese Cantonese/Toishanese, where 91.7% of individuals who speak Chinese Cantonese/Toishanese are within this cluster. Conversely, only 8.3% of individuals who speak Chinese Mandarin are within the Southern Han Chinese cluster. Furthermore, the Han Chinese cluster correlates with Chinese Mandarin, although to a lesser extent, where 66.4% of individuals who speak Chinese Mandarin fall within the Han Chinese ancestry cluster and 26.2% within the Southern Han Chinese cluster. Additionally, from Figure 3A, there are two notable clusters that do not match any of the East Asian subcontinental populations within 1000 Genomes. Projecting individuals’ self-identified race over the PCs shows that the majority of individuals in these two clusters identify as ‘Asian: Korean’ and ‘Asian: Filipino’ respectively (Supplementary Figure 5A). This pattern is similarly reflected by the self-identified preferred languages where many of these individuals speak Korean and Tagalog. This clustering not only characterizes the fine-scale genetic and ethnic diversity of ATLAS, but also emphasizes how the concepts of genetic ancestry and self-reported constructs, such as primary spoken language, can be combined to identify and label distinct genetic clusters that would not have been characterized based on a single criterion alone.

**Figure 3:**
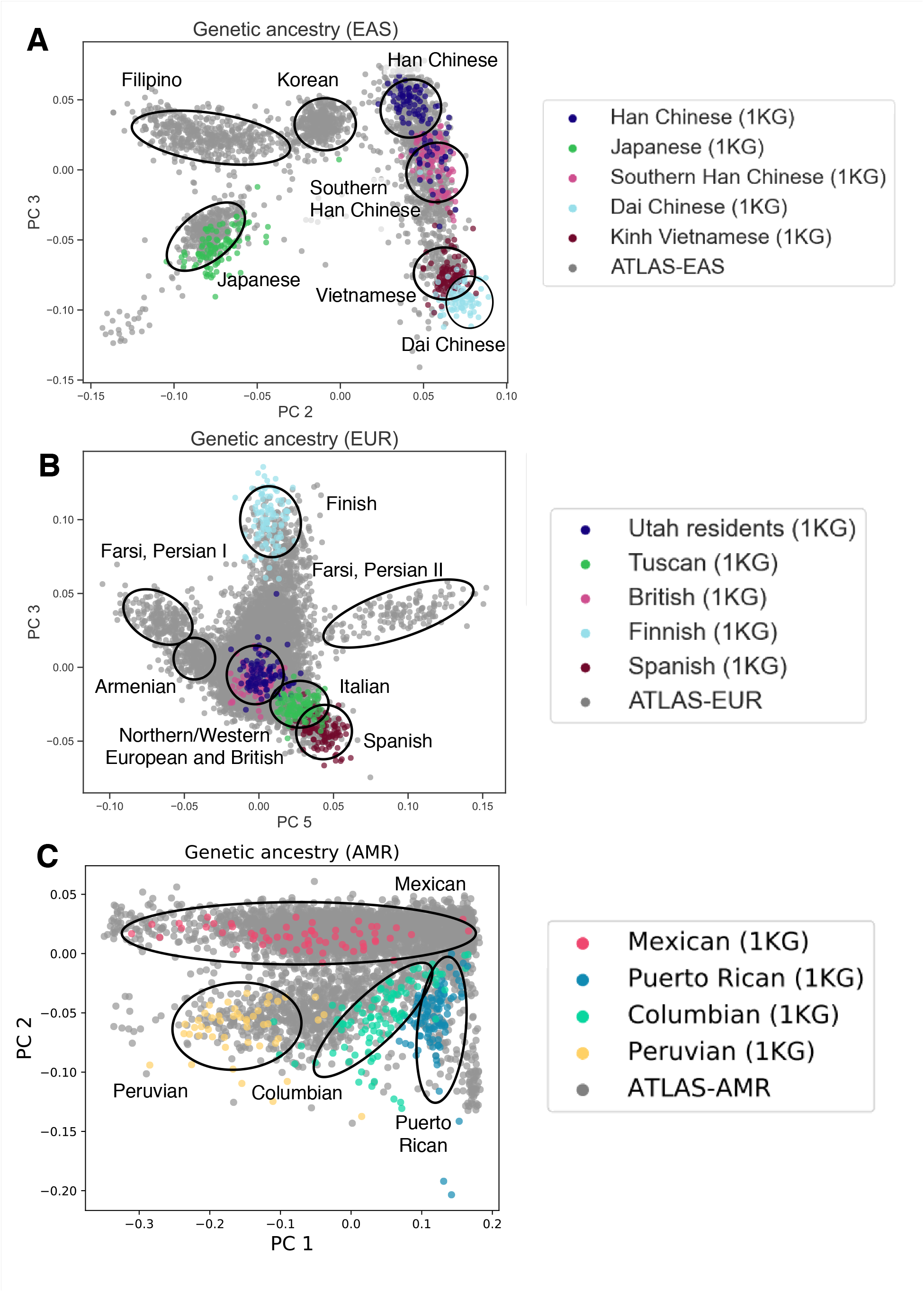
PCA of individuals with East Asian, European, and Admixed American genetic ancestry in ATLAS captures fine-scale subcontinental ancestry groupings. Principal component analysis performed separately on each continental ancestry group (East Asian, European, Admixed American) in ATLAS with corresponding subcontinental ancestry samples from 1000 Genomes. Cluster annotation labels were determined using a combination of known genetic ancestry samples from 1000 Genomes and self-reported race, ethnicity, and language information from the EHR.

Next, we identify clusters of individuals with subcontinental genetic ancestry of European descent, but due to limitations in reference panels, we were unable to describe the origins of the majority of the observed genetic variation within the ATLAS European continental ancestry cluster (Figure 3B). Comparing self-reported race and ethnicity information does not delineate any subgroups since most individuals are within the NH-WC SIRE (Supplementary Figure S6A). Instead, we overlay individuals’ self-reported preferred language over the projected PCs and observe clusters of individuals whose preferred languages are Arabic, Armenian, and Farsi/Persian; notably the primary populations that speak these languages are not present in the current 1000 Genomes reference panel (Supplementary Figure S6B). Although not definitive about ancestral origins, these results suggest that individuals in these clusters may have cultural ties and/or genetic origins relating to the Middle East. We also observe two distinct clusters of individuals who speak Farsi/Persian (labeled as ‘Farsi, Persian I’ and ‘Farsi, Persian II’), suggesting that although these groups may share cultural ties, the groups could have varying ancestral origins.

We perform a similar analysis for the Admixed American cluster of individuals. We are able to cluster individuals according to Mexican, Peruvian, Columbian, and Puerto Rican ancestry, where 66.4% of individuals are within the Mexican ancestry cluster (Figure 3C, Supplementary Figure S4). Comparing self-reported race/ethnicity and language did not reveal any additional subclusters for this population (Supplementary Figure S7A,B). However, shading the PCs by estimated genetic ancestry proportions (see Methods), we see a cline between European and Native American ancestries, demonstrating that although we cannot determine further clusters within our data, there is still substantial population substructure present (Supplementary Figure S7C,D). Corresponding analyses were also performed for the African ancestry group (Supplementary Figure S8), but clear subcontinental clusters could not be constructed. We omitted the subcontinental analysis for the South Asian ancestry group due to the small sample size.

### IBD sharing reveals communities of recent shared ancestry within ATLAS

A complementary method to principal components for inferring fine-scale ancestry is identical-by-descent (IBD) analysis [24]–[26]. Using pair-wise IBD estimates for all individuals in ATLAS and reference population information from the 1000 Genomes Project [23], Simons Genome Diversity Project [27], and Human Genome Diversity Project [28], we describe fine-scale populations based on total pairwise IBD (Figure 4; see Methods). Each subgroup is annotated according to a combination of genetic ancestry from reference populations as well as self-reported race, ethnicity, and language information. Many subgroups have similar characteristics to those defined from PCA-based clustering, such as the Filipino and Dai Chinese clusters. We can also characterize subgroups not previously identified through the previous PCA analysis. For example, PCA-based clustering was only able to distinguish clusters at the level of continental African ancestry, whereas IBD clustering identified West African, East African, and Ethiopian subgroups. In contrast, Japanese and Korean individuals form a single subgroup when estimated by the IBD clustering approach, whereas PCA-based clustering delineated these individuals into two separate groups. Note that both IBD and PCA-based clustering granularity is dependent on the clustering algorithm used and here we report at only a single level of resolution. For further discussion of PCA and IBD for fine-scale population analyses, see Belbin et al 2021[29]. Our results show that fine-scale population identification is specific to each genetic ancestry inference method, as well as how the combination of multiple methods can maximize the number of identified subgroups.

**Figure 4:**
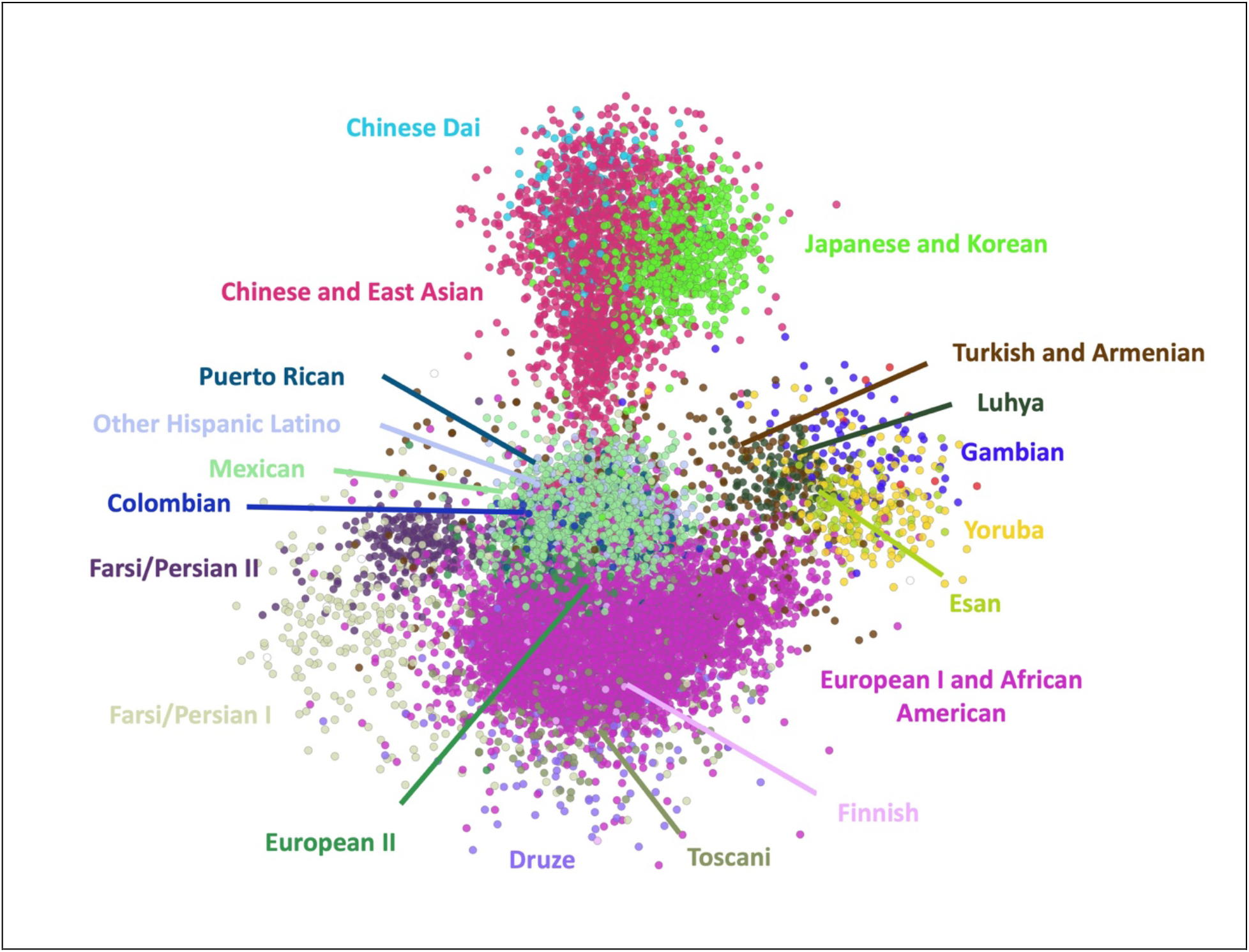
IBD sharing between ATLAS participants. InfoMap community membership is indicated by color for all communities with greater than 100 individuals (20 communities total) and individuals with a degree greater than 30. Community membership indicates elevated shared IBD within that community. Community identity is labelled adjacent to the network plot in the corresponding color.

### Admixture describes genetic variation within self-reported race/ethnicity groups

Many individuals do not fall within a single genetic ancestry cluster, but instead lie on the spectrum between multiple ancestry groups. We can characterize this variation through genetic admixture, the exchange of genetic information across two or more populations [30]. We estimate genetic ancestry proportions using k=4, 5,or 6 ancestral populations and visualize groups of individuals by SIRE (see Methods; Supplementary Figure S9). For the following analyses, we use k=4 ancestral populations where the clusters correspond to European, African, East Asian, and Native American ancestry. Among individuals in the HL-Other SIRE, the estimated average proportion of European ancestry is 49%, 6% African ancestry, and 44% Native American ancestry (Supplementary Table S6). We also observe that the HL-Other and HL-WC (White or Caucasian) SIREs have approximately the same admixture profile, where the proportion of European ancestry is 49% and 58% respectively, 6% and 5% African ancestry, and 44% and 36% Native American ancestry. However, there is also a large amount of variation within SIREs, where for example, individuals who identify as Hispanic or Latino ethnicity are estimated to have European ancestry percentages ranging from nearly 0% to almost 100%.

### Genetic ancestry groups correlate with disease prevalence within ATLAS

Understanding how disease prevalence varies across populations is integral to understanding how the interplay of genetic factors and social determinants of health contribute to disease risk. We investigate 1,330 EHR-derived phenotypes (phecodes) [31] spanning a wide range of disease categories (see Methods) and identify 1,401 total significant phecode-ancestry associations (p < 3.8e-5) across the 5 continental ancestry groups after adjusting for age and sex (Supplementary Table S7). Overall, there are 659 phenotypes that show cross-ancestry differences, where a phenotype is significantly associated with a particular ancestry group compared to the rest of the population. From this set, the highest number of phecodes are from the circulatory (N=84), endocrine/metabolic (N=74), and digestive (N=80) system-related groups. Specifically, we recapitulate many known associations such as liver and intrahepatic bile duct cancer (p=6.97e-35) within the East Asian ancestry population [32]–[34], skin cancer (p=2.02e-162) in the European ancestry population [35], [36], hereditary hemolytic anemia (p=2.4e-22) [37] and primary open-angle glaucoma (POAG) (p=5.33e-12) [38], [39] within the African ancestry population, as well as both alcoholic liver damage (p=2.0e-47) and cirrhosis of liver without mention of alcohol (p=4.84e-70) [40]–[43] in the Admixed American population (Figure 5).

**Figure 5:**
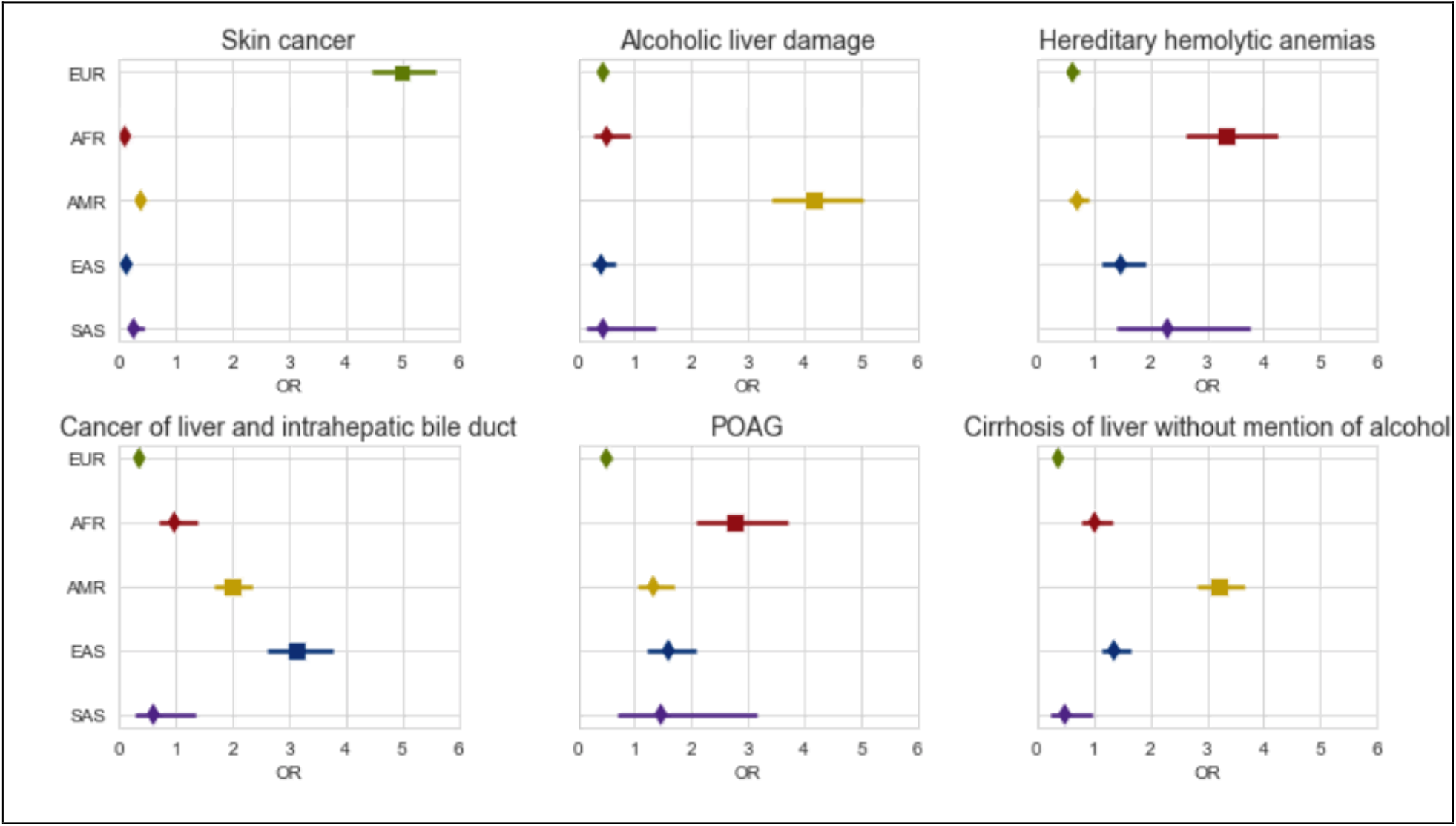
Disease associations vary across continental genetic ancestry groups in ATLAS. We show the odds ratio computed from associating each phenotype with individuals’ continental genetic ancestry in ATLAS (N=25,872) under a logistic regression model. Error bars represent 95% confidence intervals.

Next, as an example, we analyze phecodes spanning different traits where we observe a significantly higher prevalence for at least one continental ancestry group per trait. For example, we observe that the prevalence of both schizophrenia (freq=0.02, SE=0.004) and sickle cell anemia (freq=0.03, SE=0.005) have the highest prevalence in the African ancestry group in ATLAS, which is consistent with previous findings [44], [45]. We also observe substantial disease risk heterogeneity across subgroups of the same continental ancestry. We compute the prevalence for the same set of diseases across subgroups within the East Asian ancestry group (Korean, Japanese, Filipino, Chinese, and Vietnamese) in ATLAS and compare this with the aggregated East Asian ancestry group. The estimated prevalence of type 2 diabetes from the East Asian ancestry group is 0.26 (SE=0.009). However, analysis of specific subgroups shows a significant increase in the prevalence of type 2 diabetes for individuals in the Japanese (freq=0.33, SE=0.03) and Filipino (freq=0.32, SE=0.02) subgroups compared to the Chinese subgroup (freq=0.21, SE=0.01). These results indicate that genetically grouping individuals across sub-continental ancestries yield meaningful interpretation of disease risk across groups of individuals.

We also investigated disease prevalence within admixed individuals, where variation in genetic ancestry across individuals in the population allows for the correlation of disease risk with the proportion of genetic ancestry from any given continental group. Within each SIRE group, we perform an association test between the proportions of inferred ancestry estimated from ADMIXTURE [46] and each phenotype (see Methods; Supplementary Table S8). After correcting for the number of tested phenotypes, we find numerous significant phenotype-ancestry associations: 113 associations within the HL-Other SIRE, 62 within the NH-WC SIRE, and 48 within the NH-Asian SIRE. However, we do not find any significant associations within the NH-AfAm SIRE, which could be due to the smaller sample size. Across SIREs, both the top associated phenotype categories as well as the direction of the associations greatly vary. Out of the top 3 phenotype categories with the most associations in each SIRE group, only the endocrine/metabolic category is shared across all 3 tested SIREs (HL-Other, NH-WC, NH-Asian). Even within this category, looking at the statistics quantifying the association of the proportion of European ancestry with endocrine/metabolic phenotypes, there are exclusively 5 negative associations within the NH-WC group, exclusively 16 negative associations within the HL-Other group, but 8 positive associations and no negative associations within the NH-Asian group. The other top phenotype categories for each SIRE are also unique, where the HL-Other SIRE’s top categories include digestive and respiratory phenotypes, the NH-WC SIRE’s top categories include neoplasms and dermatologic phenotypes, and the NH-Asian SIRE’s top categories includes mental disorders and infectious diseases. Specifically, we find that within the HL-Other population, the overall proportion of European ancestry is significantly negatively associated (p=9.2e-14) with type 2 diabetes and the proportion of Native American ancestry is significantly positively associated (p=1.6e-13) (Figure 6A), which is consistent with previous studies [47], [48]; we additionally find a similar trend for ‘other chronic nonalcoholic liver disease’ (Figure 6B). These results suggest that not only are some disease statuses associated with SIRE and continental genetic ancestry, but the specific ancestry proportions may also correlate with disease risk.

**Figure 6:**
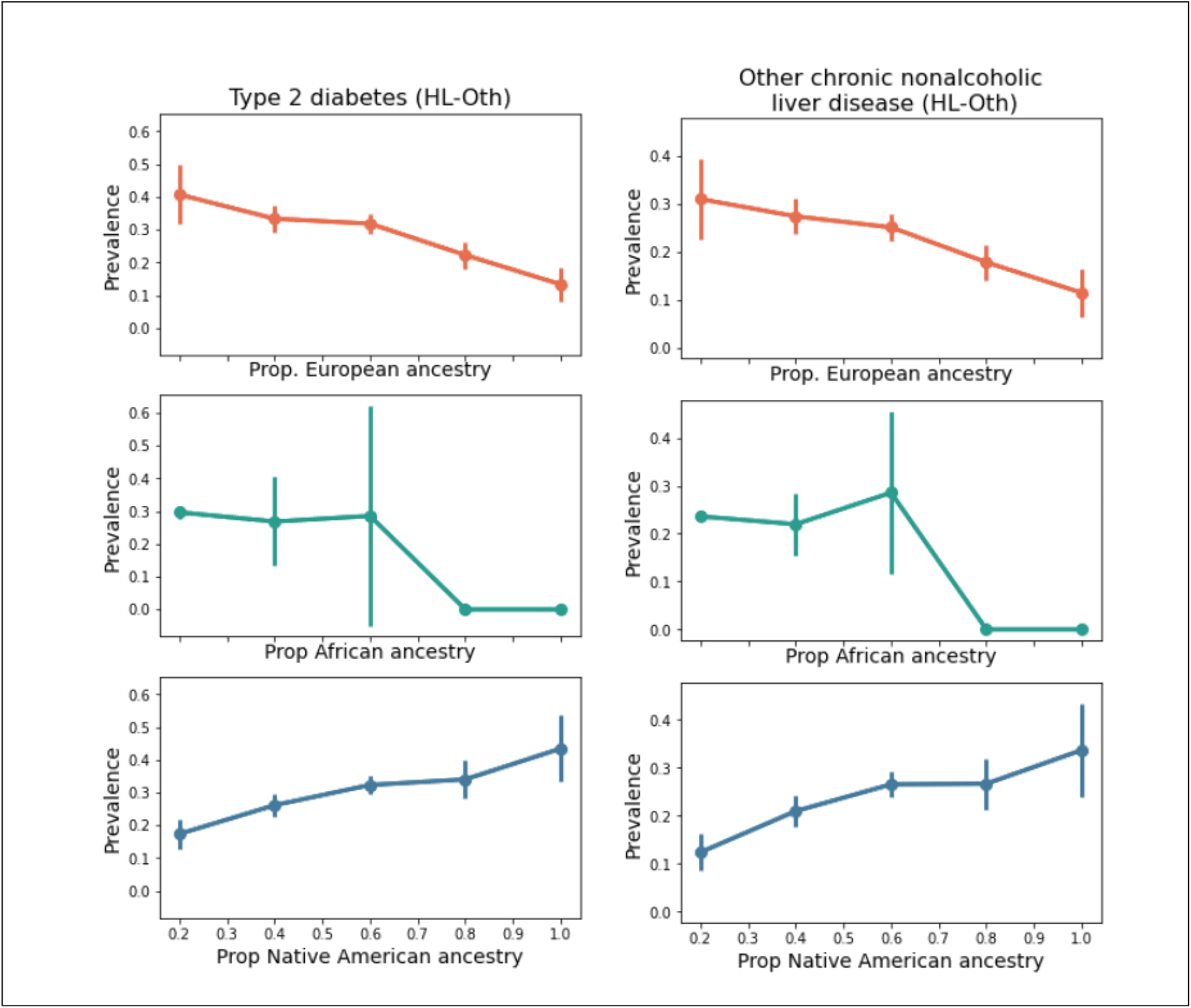
Global ancestry correlates with disease prevalence in admixed individuals. Individuals who self-report as “Hispanic/Latino – Other Race” (HL-Oth) (N=2,206) and have had a diagnosis of “Other chronic nonalcoholic liver disease” or (B) type 2 diabetes are binned by their proportions of European, African, and Native American ancestry estimated using ADMIXTURE. Bins are defined by the proportion of each ancestral population in increments of 0.20. Within each bin, we plot the prevalence of the diagnoses and provide standard errors (+/-1.96 SE) of the computed frequencies.

### Genome and phenome-wide association scans identify known risk regions and elucidates correlated phenotypes

EHR-linked biobanks also offer the opportunity of investigating genetic associations with traits across the genome. These efforts impose special challenges, such as adjusting for population stratification and cryptic relatedness in health systems that serve entire families as well as extracting phenotypes from EHR, namely due to inconsistencies in mapping diagnosis codes (ICD codes) to phenotypes and difficulties in defining appropriate controls for specific phenotypes. Here, we implemented the phecode system (v1.2) [31], [49] within a GWAS pipeline that accounts for population stratification (see Methods). As an example, we present results for phecode *571*.*5 ‘other chronic nonalcoholic liver disease’* (see Methods) across 15,439 unrelated individuals of European (EUR) ancestry and 4,472 unrelated individuals of Admixed American (AMR) ancestry (Figure 7). GWAS associations are well-calibrated for both populations (Supplementary Figure S10), with little evidence of test statistic inflation (AMR genomic inflation factor λGC = 1.01; EUR genomic inflation factor λGC = 1.02).

**Figure 7:**
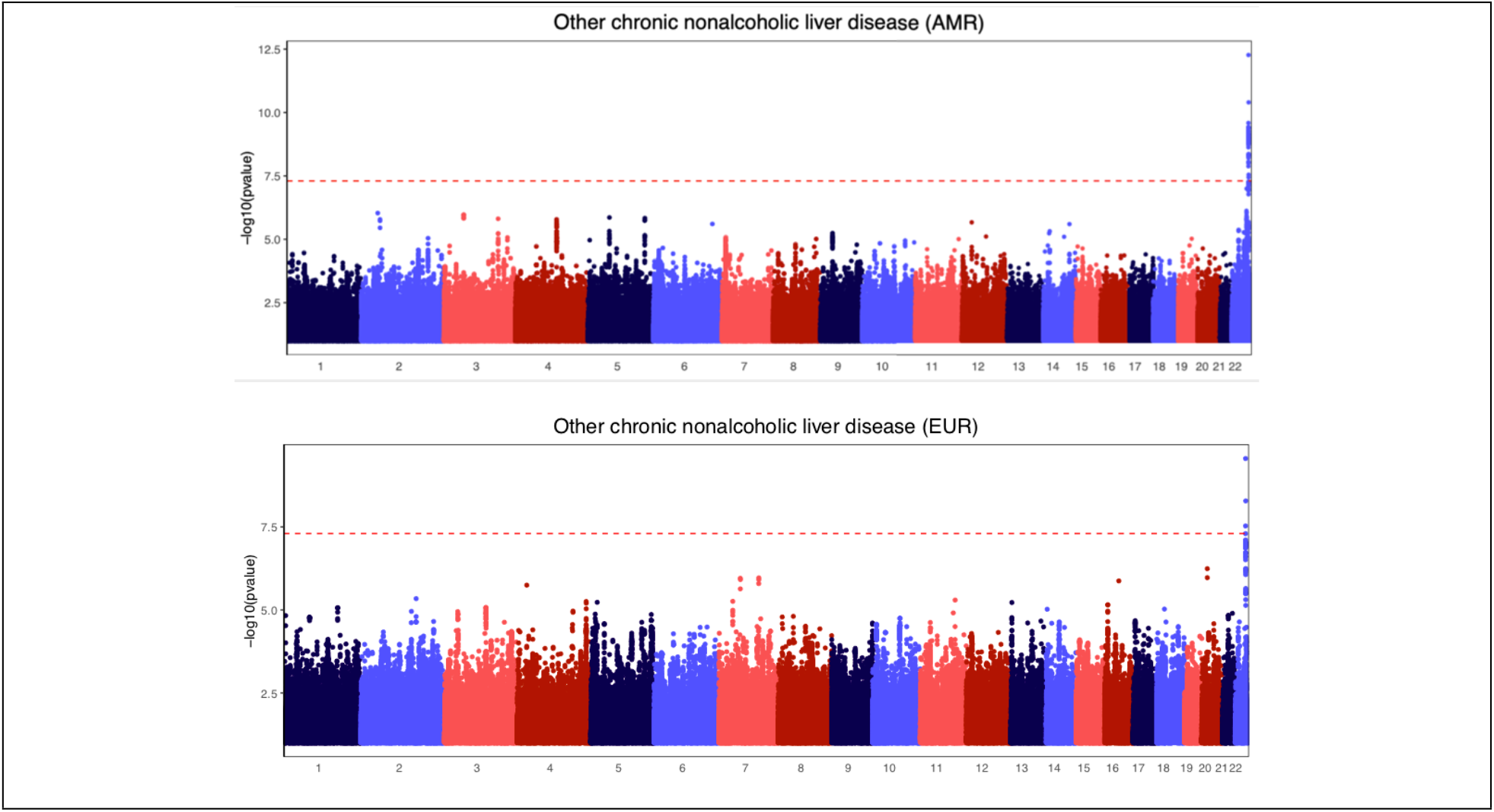
Recapitulating known associations for ‘Other chronic nonalcoholic liver disease’ GWAS in both the AMR and EUR populations in ATLAS. GWAS Manhattan plots for “Other chronic nonalcoholic liver disease” in the (A) AMR genetic ancestry group (N-Case: 919, N-Controls: 3,262) and (B) EUR genetic ancestry group (N-Case: 2,275, N-Controls: 14,155). The red dashed line denotes genome-wide significance (p<5e-08). We recapitulate a known association at the 22q13.31 locus in both populations.

In the EUR study, we find three SNPs that pass genome-wide significance (p < 5e-8) and 70 SNPs that reach significance in the AMR study (Supplementary Table S9). All genome-wide significant SNPs from both studies fall within the 22q13.31 locus, which contains the *PNPLA3* gene. This gene has been extensively studied for its role in the risk of various liver diseases such as nonalcoholic fatty liver disease [50], [51]. The lead SNP from both analyses, rs2294915, is an intronic variant in the *PNPLA3* gene and has MAF=0.45 in the AMR group but only MAF=0.24 in the EUR group. A nearby SNP, though not directly tested due to quality control filtering, is rs738409, a missense variant for PNPLA3 that has been well-documented for its role in the susceptibility of several types of liver disease [52]. Using measurements of LD from the 1000 Genomes reference panel, we find that rs2294915 is in high LD with rs738409 in the AMR analysis (R^2^ =0.94) as well as in the EUR analysis, although to a slightly lesser extent (R^2^ =0.85) [53].

Next, we leverage GWAS for all existing phecodes to investigate the association of the lead variant, rs2294915, across all 1,330 EHR-derived phenotypes (i.e. a phenome-wide association study: PheWAS). After adjusting for both genome-wide significance and the number of phenotypes (p < 3.8e-11), we find that only the liver-related phenotypes within the AMR study reach significance (Figure 8). Additionally, multiple neoplastic and neurological phenotypes, which are comorbidities with severe liver disease [50], [54]–[56], are nominally significant only in the AMR study after adjusting for the number of tested phenotypes (p < 3.8e-5). These findings suggest possible differential genetic architecture across these two populations, as well as variation even at the phenotype level, reflecting possible genetic or environmental modifiers of important comorbidities.

**Figure 8:**
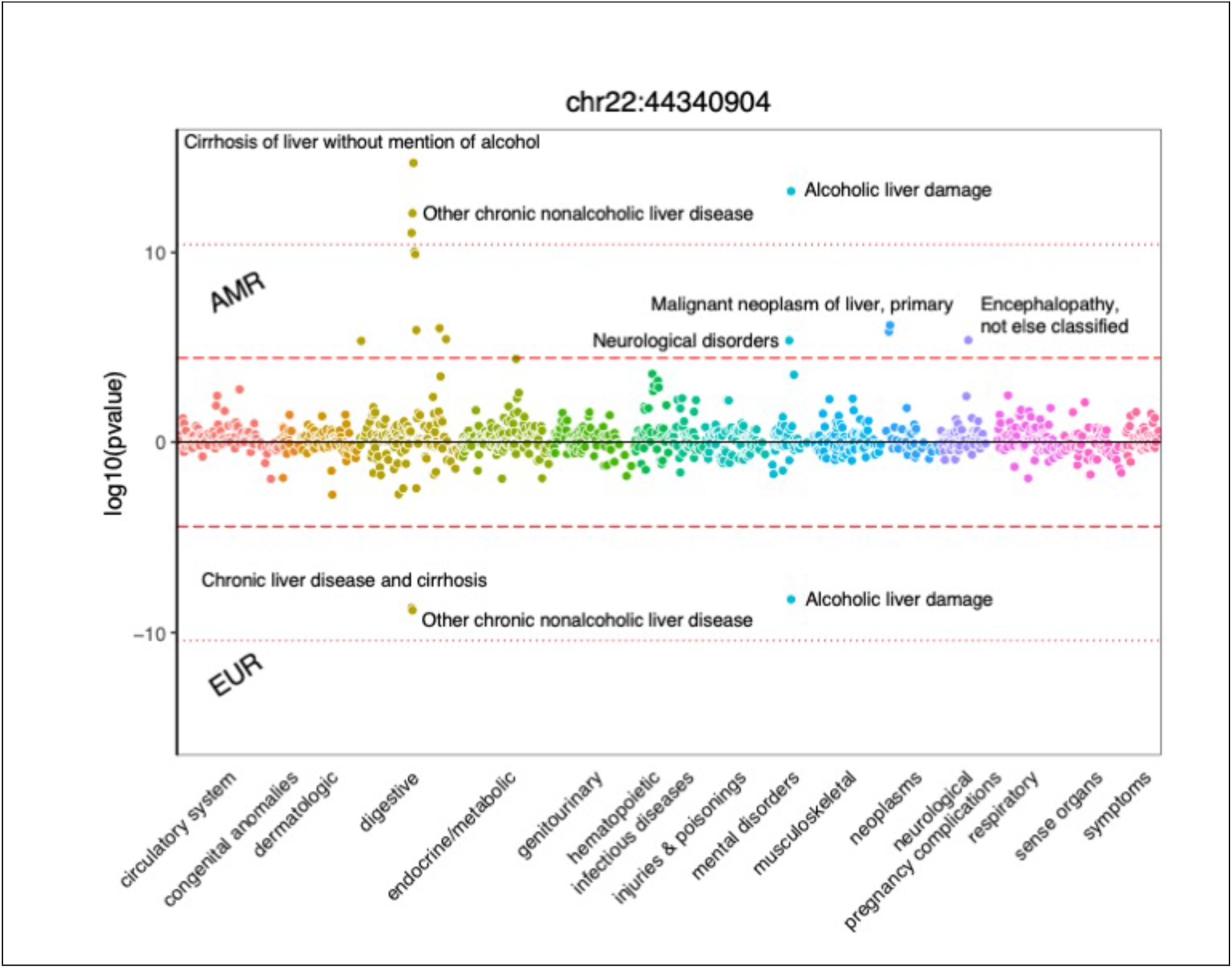
Identifying correlated phenotypes at rs2294915 in both the Admixed American and European populations in ATLAS. PheWAS plot at rs2294915 for the Admixed American (top) and European (bottom) ancestry groups across 1,330 phecode phenotypes. The red dotted line denotes p=3.8e-11, the significance threshold after adjusting the genome-wide significance threshold for 1,330 tested phenotypes. The red dashed line denotes p=3.8e-5, the significance threshold after correcting for only the 1330 tested phenotypes.

## Discussion

In this work, we introduce the ATLAS Community Health Initiative, a biobank embedded within the UCLA Health medical system comprising of de-identified EHR-linked genomic data from a diverse patient population. The UCLA Health system serves Los Angeles County, leading to a study population of great demographic, genetic, and phenotypic diversity. We investigate ancestry both on the continental as well as the subcontinental population level and find that genetic ancestry and self-reported demographic information yield distinct subpopulations in the ATLAS biobank. We present a collection of results cataloguing the associations between genetic ancestry and EHR-derived phenotype where we find that disease status is not only associated with continental genetic ancestry but also associated with the specific admixture profile describing an individual. We use multi-ethnic pipelines to recapitulate known associations for chronic nonalcoholic liver disease at the 22q13.31 locus and perform a phenome-wide association study at the lead SNP, where we find associations with neurological and neoplastic phenotypes exclusively in the Admixed American ancestry group. As the sample size increases, the ATLAS Community Health Initiative will enable rigorous genetic and epidemiological studies to further understand the role of genetic ancestry in disease etiology, with a specific aim to accelerate genomic medicine in diverse populations. Already, the ATLAS biobank accounted for 73.4% of the Admixed American samples utilized in the primary analysis from the COVID-19 Host Genetics Initiative [57].

As the field moves forward with increased collaboration between the genetics and healthcare communities, it is of utmost importance to also be aware of potential pitfalls that may occur when translating research findings into actual clinical populations. Currently, many clinical protocols are deeply ingrained with racial bias, no matter how benign the original goal was intended[58]–[62]. Many of these flawed policies stemmed from erroneously linking race, a social rather than biological construct, with disease risk despite not presenting any biological justification. Although race and genetic ancestry are correlated [63], [64], our work shows that populations constructed from these two concepts are not analogous. We encourage protocol decisions that are rooted in actual biological phenomena, such as genetic markers, providing transparent, immutable criteria. For example, Benign Ethnic Neutropenia (BEN) is observed predominantly in African Americans, but specifically is strongly associated with the variant at rs2814778 [65], [66]. Recent studies have suggested that genotype screening at rs2814778 could aid in the interpretation of neutropenia in African Americans and avoid unnecessary invasive procedures as well as lead to an increase of the inclusion of these individuals to various treatments [67]. Additionally, the Kidney Donor Risk Index (KDRI) equation uses race as a risk factor [68], but it has been recently proposed to use the presence of *APOL1* variants as a factor instead [69]. Discovered after the creation of the KDRI, the presence of these variants was shown to be associated with shorter allograft longevity [70]. Despite this finding, the original KDRI score is still commonly used. In order to remedy and not perpetuate current healthcare inequalities, we underscore the importance of favoring transparent clinical protocols with clear biological justification instead of race-adjusted formulas that leverage convenience at the expense of potential inequities.

There are various limitations within our study, and we describe a few of these in detail as follows. First, the phenotypes are based on ICD codes, and due to the nature of billing codes, this form of labeling does not constitute a formal patient diagnosis and may contain individuals who do not have the specific disease. This uncertainty in phenotyping likely limits the power of our study to find disease associations. For the further investigation into specific phenotypes, we recommend refining each phenotype definition based on additional disease-specific factors and metrics. Additionally, although ICD codes are an international standard, there are still deviations between different institutions in how specific diagnoses are recorded. This adds further heterogeneity in phenotyping and could present future challenges when replicating studies or porting algorithms to other institutions. Second, due to the de-identified nature of the data, we lack information that could help us better describe the fine-scale population groups. For example, location of birth, zip code, and family history has been shown to be useful descriptors for determining subgroups of genetic ancestry [29]. This geographic information could also be used as a proxy for various environmental exposures such as pollution. Additional socioeconomic information, such as income and availability of health insurance, could likely account for a portion of observed associations as well as provide more insight into the socioeconomic determinants of health. Third, our findings within the African and South Asian ancestry populations are limited due to the smaller sample sizes. As sample sizes increase, we hope to further refine population substructure within these initial continental ancestry groups and have the power to detect novel disease associations that have previously been mired by lack of statistical power.

We conclude by discussing directions for future work. Although we investigate admixed populations, such as African American and Hispanic/Latino populations, admixed individuals who do not fall under these groups are excluded from downstream analyses due to concerns over population structure. In the future, we hope to incorporate methods and pipelines that properly control for population structure in all types of admixed populations. Additionally, we plan to compute polygenic risk scores (PRS) across all 5 continental ancestry groups. PRS has already shown modest clinical utility for diseases such as breast cancer [71] and cardiovascular disease [72], but has proven difficult to perform accurate predictions across populations [13]. The genetic diversity within the ATLAS Community Health Initiative biobank partnered with the longitudinal clinical data provides a unique resource to further explore the role of ancestry in PRS prediction. Furthermore, as the size of the biobank grows and more data is collected over time, we hope to explore even more individualized health solutions and interventions.

## Methods

### Study population

The UCLA Health System includes two hospitals (520 and 281 inpatient beds) and 210 primary and specialty outpatient locations predominantly located in Los Angeles County. The UCLA Data Discovery Repository (DDR) contains de-identified patient electronic health records (EHR) that have been collected since March 2^nd^, 2013, under the auspices of the UCLA Health Office of Health Informatics Analytics and the UCLA Institute of Precision Health. Currently, the DDR contains longitudinal records for more than 1.5 million patients, including basic patient information (height, weight, gender), diagnosis codes, laboratory tests, medications, prescriptions, hospital admissions, and procedures. The UCLA ATLAS Community Health Initiative includes the EHR-linked biobank within the UCLA Health System. Currently, there are more than 27,000 genotyped participants with their de-identified EHR linked through the DDR. Patients’ participation is voluntary and their privacy is protected by de-identifying the samples.

### Self-reported demographic information

Participants self-report their race and ethnicity via two distinct fields where there are multiple-choice fields for race and ethnicity (see Supplementary Table S1, S2 for full list). Only one selection from each category can be chosen as a patient’s primary race and ethnicity. We group together race/ethnicity pairings to form 21 ‘self-reported race/ethnicity’ (SIRE) groupings (see Supplementary Table S3). Patients also report their ‘Preferred Language’ from multiple-choice fields.

### Genotype quality control

Bio-samples collected from the UCLA ATLAS Community Health Initiative in the form of blood samples, were de-identified and then processed for DNA extraction and genotyping. ATLAS participants (N= 26,439) were genotyped on a custom Illumina Global Screening (GSA) array [17] that included a standard GWAS backbone and an additional set of pathogenic variants selected from ClinVar [73]. We filtered out poor quality markers by removing variants with >5% missingness (M= 9,318 variants removed) and then removed strand-ambiguous SNPs (M= 7,686). We excluded samples with missingness >5% (N=3 individuals removed) and kept one individual from each set of twins or duplicated individuals (N=22 individuals removed). All quality control steps were conducted using PLINK 1.9 [74] and duplicate individuals/twins were determined using KING [75]. These steps resulted in a total of 673,148 variants and 26,414 individuals.

### Genetic relatedness inference

With the M=673,148 variants that passed quality control, we used KING [75] to compute pairwise kinship coefficients to determine family relationships. We identified a set of unrelated individuals (N=25,842) where individuals with kinship coefficient <0.0884 were included (‘king --unrelated’). Additionally, we identified 22 duplicate individuals or twins, 213 parent-offspring relatives, and 117 first-degree relatives. This level of relatedness is expected since families will often be members of the same healthcare system.

### Genotype imputation

After performing array-level genotype quality control, the PLINK-formatted genotypes are converted to VCF format and uploaded to the Michigan Imputation Sever [76]. On a variant level, the server removes the variant if it is not an A, C, G, T allele, monomorphic, a duplicate, an allele mismatch between the reference panel and provided data, an insertion-deletion, or if the SNP call rate is less than 90%. The server will additionally correct for any necessary strand flips or allele switches needed to match the reference panel. The server additionally phases the data using Eagle v2.4 [77] and imputation is performed against the TOPMed Freeze5 imputation panel [78] using minimac4 [79]. In summary, the explicit parameters used on the server are “TOPMed Freeze5” for the reference panel, “GRCh38/hg38” for the array build, “off” for the rsq filter, “Eagle v2.4” for phasing, no QC frequency check, and “quality control & imputation” for the mode. After we filtered by R2>0.90 and MAF>1%, the final set of variants contained M=7,973,837 sites.

### Genetic ancestry PCA analysis

We limited analyses to unrelated individuals (2nd degree relatives) and performed the ancestry-related PCA analyses on only the typed data. Genotypes were filtered by Mendel error rate (‘plink --me 1 1 –set-me-missing’), founders (‘--filter-founders’), minor allele frequency (‘–maf 0.15’), genotype missing call rate (‘--geno 0.05’), and Hardy-Weinberg equilibrium test p-value (‘–hwe 0.001’). Genotypes were then merged with the 1000 Genomes phase 3 dataset [23]. LD pruning was then performed on the merged dataset (‘--indep 200 5 1.15 --indep-pairwise 100 5 0.1’). We computed the top 10 PCs using the FlashPCA software [80] with all default settings.

We use the superpopulations from the 1000 Genomes dataset to define continental genetic ancestry groups (European, African, East Asian, South Asian). Ancestry clusters were determined by visually defining PC thresholds based on the labeled individuals from the 1000 Genomes dataset. The European group was determined by PC thresholds with PC1 and PC2 (Supplementary Figure S1A), the African group with PC1 and PC2 (Supplementary Figure S1B), the Admixed American group with PC2 and PC3 (Supplementary Figure S1C), the East Asian group with PC1 and PC2 (Supplementary Figure S1D), and the South Asian group with PC4 and PC5 (Supplementary Figure S1E). ATLAS participants that fell within the set of thresholds for each ancestry group were assigned to that ancestry. Individuals who were classified into multiple ancestry groups or individuals that could not be classified into any of the ancestry groups were given an ancestry label of ‘Admixed or other ancestry’.

### Subcontinental PCA analysis

We ran PCA on individuals within the East Asian continental genetic ancestry group (N=2,242) and the individuals from the East Asian superpopulation within 1000 Genomes using FlashPCA. We define clusters of hypothesized Chinese, Vietnamese, Korean, Japanese, and Filipino ancestry. Ancestry groups were determined by visually defining PC thresholds based on the self-reported race information of the ATLAS participants: Korean ancestry based on PC2 (Supplementary Figure S3A), Chinese ancestry based on PC1 (Supplementary Figure S3B), Vietnamese ancestry based on PC1 (Supplementary Figure S3C), Filipino ancestry based on PC1 (Supplementary Figure S3D), and Japanese ancestry based on PC1 and PC3 (Supplementary Figure S3E, S3F).

We additionally ran PCA on individuals from the initial Admixed American ancestry cluster in ATLAS and individuals of Mexican (MXL), Puerto Rican (PUR), Columbian (CLM), and Peruvian (PEL) ancestry from 1000 Genomes. The Mexican ancestry cluster in ATLAS is described by PC1 (Supplementary Figure 4A) and the Puerto Rican cluster is determined by PCs 1, 2, and 8 (Supplementary Figure 4B, C, D). Explicit thresholds could not confidently be drawn for the Columbian and Peruvian ancestry clusters in ATLAS. Additionally, we did not compute explicit PC thresholds for the European subcontinental clusters.

### GWAS quality control per ancestry

We limited our analyses to N=25,842 unrelated individuals and then performed additional quality control steps within each continental ancestry groups for GWAS (European, African, Admixed American, East Asian, South Asian). SNPs that violated Hardy-Weinberg equilibrium (HWE) with p<1e-12 were excluded. Individuals with a heterozygosity rate that surpassed +/-3 standard deviations from the ancestry-specific mean were also excluded. Analyses were restricted to common SNPs per ancestry group where MAF>1%.

### IBD Calling

For IBD calling, an interim version of the ATLAS data comprising of 24,318 individuals was used. ATLAS data was merged with the 1000 Genome Project [23], the Simons Genome Diversity Project [27], and the Human Genome Diversity Project [28]. In total, 418,195 SNPs were kept for IBD analysis after filtering by missingness >=10% and MAF>1%. The merged dataset was then statistically phased using Shapeit4 [81]. IBD was called using iLASH using default parameters [82]. For downstream analysis, IBD segments were summed between individuals to create an adjacency matrix, where each row represented a pair of individuals, and each column represented the total genome-wide IBD between those two individuals. Using KING [75], the adjacency matrix was filtered to remove rows representing individuals who were third degree relatives or closer. Communities are annotated using the presence of reference individuals in a cluster and EHR characteristics, such as preferred language and self-reported race/ethnicity.

### Genetic admixture analysis

We inferred the proportion of genetic ancestry by using the AMIXTURE software [46] under the unsupervised clustering mode with the number of clusters k=4, 5, 6. For each SIRE, we compare the ancestry proportions from the clusters. For k=4, we assign the cluster with the majority of NH-WC individuals as European ancestry, the cluster with the majority of NH-AfAm individuals as African ancestry, the cluster with the majority of NH-Asian individuals as East Asian ancestry, and the cluster with the highest number of HL-Other and HL-WC individuals as Native American ancestry.

### Phecodes

We aggregated billing (ICD9/ICD10) codes into more clinically informative groupings known as phecodes [31]. We derived phecodes from ICD codes in the EHR using mappings described in the PheWAS catalogue (Phecode Map 1.2) [83]. Using phecodes to define case/control phenotypes, we treated individuals with the occurrence of a specific phecode as a case and a control otherwise. We restricted our analyses to phecodes that had >100 cases present in ATLAS, yielding a total of 1330 phenotypes.

### Association between phecodes and genetic ancestry

We performed a marginal association between each phecode and continental genetic ancestry group under a logistic regression model while also adjusting for age and sex. Statistical significance was determined after correcting for the number of tested phecodes (p<3.8e-5).

### Association between genetic admixture proportions and phecodes

We perform a marginal regression between each of the ancestry proportions estimated from ADMIXTURE where k=4 (European, African, East Asian, and Native American ancestry) and 1,300 EHR-derived phenotypes (phecodes) within each of the 7 ATLAS SIRE groups groups (NH-WC, NH-AfAm, HL-Other, HL-WC, NH-Asian, NH-PI, NH-AmIn). Additional details on computing admixture proportions can be found under the section *Genetic admixture analysis*. We additionally adjust for age and sex in the regression. Only traits with >10 cases per SIRE were tested. Significance is determined after adjusting for the number of tested phenotypes (p<3.8e-5).

### GWAS for ‘Other chronic nonalcoholic liver disease’

We performed an association between all imputed autosomal variants and ‘Other chronic nonalcoholic liver disease’ within the European (N-Case: 2,275, N-Controls: 14,155) and Admixed American (N-Case=919; N-Controls=3,262) continental ancestry groups. Using PLINK 1.9, we performed a marginal association test at each SNP using a logistic regression model (‘plink --logistic beta’) where we adjusted for age, sex, and PCs 1-10. Quantile-quantile plots and genomic inflation factors (EUR λ_GC_ = 1.02; AMR λ_GC_ = 1.01) provide evidence that both analyses are well-calibrated (Supplementary Figure S10).

### PheWAS

We performed an association between each typed SNP and 1330 phecodes. Due to the number of tests, we only perform associations at genotyped SNPs. To determine significance, we used a stringent threshold that corrects for both the number of tested phenotypes as well as genome-wide significance (p<3.8e-11) and a less stringent threshold that only corrects for genome-wide significance (p<5e-08).

## Data Availability

Individual electronic health record data and genomic data are not publicly available due to patient confidentiality and security concerns.

## Notes

### Competing Interest Statement

The authors have declared no competing interest.

### Funding Statement

No external funding was received.

### Author Declarations

Patient Recruitment and Sample Collection for Precision Health Activities at UCLA is an approved study by the UCLA Institutional Review Board (UCLA IRB). IRB#17-001013.

## References

[1] R. Li, Y. Chen, M. D. Ritchie, and J. H. Moore, “Electronic health records and polygenic risk scores for predicting disease risk,” Nat. Rev. Genet., vol. 21, no. 8, pp. 493–502, Aug. 2020.

[2] T. J. Morley et al., “Phenotypic signatures in clinical data enable systematic identification of patients for genetic testing,” Nature Medicine 2021 27:6, vol. 27, no. 6, pp. 1097–1104, Jun. 2021, doi: 10.1038/s41591-021-01356-z.

[3] L. Bastarache et al., “Phenotype risk scores identify patients with unrecognized mendelian disease patterns,” Science, vol. 359, no. 6381, pp. 1233–1239, Mar. 2018, doi: 10.1126/SCIENCE.AAL4043.

[4] N. S. Abul-Husn and E. E. Kenny, “Leading Edge Perspective Personalized Medicine and the Power of Electronic Health Records,” Cell, vol. 177, pp. 58–69, 2019, doi: 10.1016/j.cell.2019.02.039.

[5] C. K. Svensson, “Representation of American blacks in clinical trials of new drugs,” JAMA, vol. 261, no. 2, pp. 263–265, Jan. 1989.

[6] V. H. Murthy, H. M. Krumholz, and C. P. Gross, “Participation in cancer clinical trials: race-, sex-, and age-based disparities,” JAMA, vol. 291, no. 22, pp. 2720–2726, Jun. 2004.

[7] A. B. Popejoy and S. M. Fullerton, “Genomics is failing on diversity,” Nature, vol. 538, no. 7624, pp. 161–164, Oct. 2016.

[8] G. Sirugo, S. M. Williams, and S. A. Tishkoff, “The Missing Diversity in Human Genetic Studies,” Cell, vol. 177, no. 1, pp. 26–31, Mar. 2019.

[9] X. Guo, E. Vittinghoff, J. E. Olgin, G. M. Marcus, and M. J. Pletcher, “Volunteer Participation in the Health eHeart Study: A Comparison with the US Population,” Sci. Rep., vol. 7, no. 1, p. 1956, May 2017.

[10] E. M. Rencsok et al., “Diversity of Enrollment in Prostate Cancer Clinical Trials: Current Status and Future Directions,” Cancer Epidemiol. Biomarkers Prev., vol. 29, no. 7, pp. 1374–1380, Jul. 2020.

[11] L. E. Flores et al., “Assessment of the Inclusion of Racial/Ethnic Minority, Female, and Older Individuals in Vaccine Clinical Trials,” JAMA Netw Open, vol. 4, no. 2, p. e2037640, Feb. 2021.

[12] A. S. Adamson and A. Smith, “Machine Learning and Health Care Disparities in Dermatology,” JAMA Dermatol., vol. 154, no. 11, pp. 1247–1248, Nov. 2018.

[13] A. R. Martin, M. Kanai, Y. Kamatani, Y. Okada, B. M. Neale, and M. J. Daly, “Clinical use of current polygenic risk scores may exacerbate health disparities,” Nat. Genet., vol. 51, no. 4, pp. 584–591, Apr. 2019.

[14] K. A. Schulman et al., “The effect of race and sex on physicians’ recommendations for cardiac catheterization,” N. Engl. J. Med., vol. 340, no. 8, pp. 618–626, Feb. 1999.

[15] D. Curtis, “Polygenic risk score for schizophrenia is more strongly associated with ancestry than with schizophrenia,” Psychiatr. Genet., vol. 28, no. 5, pp. 85–89, Oct. 2018.

[16] United States Census Bureau, “QuickFacts: Los Angeles city, California.” 2020.

[17] “Infinium Global Screening Array-24 Kit | Population-scale genetics.” https://www.illumina.com/products/by-type/microarray-kits/infinium-global-screening.html (accessed Aug. 09, 2021).

[18] L. N. Borrell, “Racial identity among Hispanics: implications for health and well-being,” Am. J. Public Health, vol. 95, no. 3, pp. 379–381, Mar. 2005.

[19] D. A. Vyas, L. G. Eisenstein, and D. S. Jones, “Hidden in Plain Sight — Reconsidering the Use of Race Correction in Clinical Algorithms,” https://doi.org/10.1056/NEJMms2004740, vol. 383, no. 9, pp. 874–882, Jun. 2020, doi: 10.1056/NEJMMS2004740.

[20] L. N. Borrell et al., “Race and Genetic Ancestry in Medicine — A Time for Reckoning with Racism,” https://doi.org/10.1056/NEJMms2029562, vol. 384, no. 5, pp. 474–480, Jan. 2021, doi: 10.1056/NEJMMS2029562.

[21] I. T. Jolliffe, “Principal Component Analysis and Factor Analysis,” in Principal Component Analysis, I. T. Jolliffe, Ed. New York, NY: Springer New York, 1986, pp. 115–128.

[22] J. Novembre et al., “Genes mirror geography within Europe,” Nature, vol. 456, no. 7218, pp. 98–101, Nov. 2008.

[23] 1000 Genomes Project Consortium et al., “A global reference for human genetic variation,” Nature, vol. 526, no. 7571, pp. 68–74, Oct. 2015.

[24] S. Carmi, P. F. Palamara, V. Vacic, T. Lencz, A. Darvasi, and I. Pe’er, “The Variance of Identity-by-Descent Sharing in the Wright–Fisher Model,” Genetics, vol. 193, no. 3, pp. 911–928, Mar. 2013, doi: 10.1534/GENETICS.112.147215.

[25] Y. Erlich, T. Shor, I. Pe’er, and S. Carmi, “Identity inference of genomic data using long-range familial searches,” Science, vol. 362, no. 6415, pp. 690–694, Nov. 2018, doi: 10.1126/SCIENCE.AAU4832.

[26] P. F. Palamara, T. Lencz, A. Darvasi, and I. Pe’er, “Length Distributions of Identity by Descent Reveal Fine-Scale Demographic History,” The American Journal of Human Genetics, vol. 91, no. 5, pp. 809–822, Nov. 2012, doi: 10.1016/J.AJHG.2012.08.030.

[27] S. Mallick et al., “The Simons Genome Diversity Project: 300 genomes from 142 diverse populations,” Nature 2016 538:7624, vol. 538, no. 7624, pp. 201–206, Sep. 2016, doi: 10.1038/nature18964.

[28] A. Bergström et al., “Insights into human genetic variation and population history from 929 diverse genomes,” Science, vol. 367, no. 6484, Mar. 2020, doi: 10.1126/SCIENCE.AAY5012.

[29] G. M. Belbin et al., “Toward a fine-scale population health monitoring system,” Cell, vol. 184, no. 8, pp. 2068-2083.e11, Apr. 2021, doi: 10.1016/J.CELL.2021.03.034.

[30] G. Hellenthal et al., “A genetic atlas of human admixture history,” Science, vol. 343, no. 6172, pp. 747–751, Feb. 2014.

[31] J. C. Denny et al., “PheWAS: demonstrating the feasibility of a phenome-wide scan to discover gene-disease associations,” Bioinformatics, vol. 26, no. 9, pp. 1205–1210, May 2010.

[32] E. T. Chang et al., “The burden of liver cancer in Asians and Pacific Islanders in the Greater San Francisco Bay Area, 1990 through 2004,” Cancer, vol. 109, no. 10, p. 2100, May 2007, doi: 10.1002/CNCR.22642.

[33] C. MS, “Cancer health disparities among Asian Americans: what we do and what we need to do,” Cancer, vol. 104, no. 12 Suppl, pp. 2895–2902, Dec. 2005, doi: 10.1002/CNCR.21501.

[34] “|Cancer Statistics Review, 1975-2018 - SEER Statistics.” https://seer.cancer.gov/csr/1975_2018/ (accessed Sep. 08, 2021).

[35] R. L. Siegel, K. D. Miller, H. E. Fuchs, and A. Jemal, “Cancer Statistics, 2021,” CA: A Cancer Journal for Clinicians, vol. 71, no. 1, pp. 7–33, Jan. 2021, doi: 10.3322/CAAC.21654.

[36] R. HW, W. MA, F. SR, and C. BM, “Incidence Estimate of Nonmelanoma Skin Cancer (Keratinocyte Carcinomas) in the U.S. Population, 2012,” JAMA dermatology, vol. 151, no. 10, pp. 1081–1086, Oct. 2015, doi: 10.1001/JAMADERMATOL.2015.1187.

[37] “Data & Statistics on Sickle Cell Disease | CDC.” https://www.cdc.gov/ncbddd/sicklecell/data.html (accessed Sep. 08, 2021).

[38] Y. C. Tham, X. Li, T. Y. Wong, H. A. Quigley, T. Aung, and C. Y. Cheng, “Global Prevalence of Glaucoma and Projections of Glaucoma Burden through 2040: A Systematic Review and Meta-Analysis,” Ophthalmology, vol. 121, no. 11, pp. 2081–2090, Nov. 2014, doi: 10.1016/J.OPHTHA.2014.05.013.

[39] N. Zhang, J. Wang, Y. Li, and B. Jiang, “Prevalence of primary open angle glaucoma in the last 20 years: a meta-analysis and systematic review,” Scientific Reports 2021 11:1, vol. 11, no. 1, pp. 1–12, Jul. 2021, doi: 10.1038/s41598-021-92971-w.

[40] M. Lazo et al., “Prevalence of Nonalcoholic Fatty Liver Disease in the United States: The Third National Health and Nutrition Examination Survey, 1988–1994,” American Journal of Epidemiology, vol. 178, no. 1, pp. 38–45, Jul. 2013, doi: 10.1093/AJE/KWS448.

[41] K. Bambha et al., “Ethnicity and nonalcoholic fatty liver disease,” Hepatology, vol. 55, no. 3, pp. 769–780, Mar. 2012, doi: 10.1002/HEP.24726.

[42] M. I. Goran and E. E. Ventura, “Genetic predisposition and increasing dietary fructose exposure: The perfect storm for fatty liver disease in Hispanics in the U.S.,” Digestive and Liver Disease, vol. 44, no. 9, pp. 711–713, Sep. 2012, doi: 10.1016/J.DLD.2012.04.009.

[43] A. F. Carrion, R. Ghanta, O. Carrasquillo, and P. Martin, “Chronic Liver Disease in the Hispanic Population of the United States,” Clinical gastroenterology and hepatology : the official clinical practice journal of the American Gastroenterological Association, vol. 9, no. 10, p. 834, Oct. 2011, doi: 10.1016/J.CGH.2011.04.027.

[44] A. Barnes, “Race, schizophrenia, and admission to state psychiatric hospitals,” Adm. Policy Ment. Health, vol. 31, no. 3, pp. 241–252, Jan. 2004.

[45] T. Somervaille, “Disorders of Hemoglobin: Genetics, Pathophysiology, and Clinical Management,” J. R. Soc. Med., vol. 94, no. 11, p. 602, Nov. 2001.

[46] D. H. Alexander, J. Novembre, and K. Lange, “Fast model-based estimation of ancestry in unrelated individuals,” Genome Res., vol. 19, no. 9, pp. 1655–1664, Sep. 2009.

[47] C.-Y. Cheng et al., “African ancestry and its correlation to type 2 diabetes in African Americans: a genetic admixture analysis in three U.S. population cohorts,” PLoS One, vol. 7, no. 3, p. e32840, Mar. 2012.

[48] A. T. Chande et al., “Influence of genetic ancestry and socioeconomic status on type 2 diabetes in the diverse Colombian populations of Chocó and Antioquia,” Sci. Rep., vol. 7, no. 1, p. 17127, Dec. 2017.

[49] J. C. Denny et al., “Systematic comparison of phenome-wide association study of electronic medical record data and genome-wide association study data,” Nature Biotechnology 2013 31:12, vol. 31, no. 12, pp. 1102–1111, Dec. 2013, doi: 10.1038/nbt.2749.

[50] L. E. Wagenknecht et al., “Association of PNPLA3 with non-alcoholic fatty liver disease in a minority cohort: the Insulin Resistance Atherosclerosis Family Study,” Liver Int., vol. 31, no. 3, pp. 412–416, Mar. 2011.

[51] E. Trépo, S. Romeo, J. Zucman-Rossi, and P. Nahon, “PNPLA3 gene in liver diseases,” J. Hepatol., vol. 65, no. 2, pp. 399–412, Aug. 2016.

[52] J.-H. Shen, Y.-L. Li, D. Li, L. Wang Ning-Ning and Jing, and Y.-H. Huang, “The rs738409 (I148M) variant of the PNPLA3 gene and cirrhosis: a meta-analysis,” J. Lipid Res., vol. 56, no. 1, pp. 167–175, Jan. 2015.

[53] M. MJ and C. SJ, “LDlink: a web-based application for exploring population-specific haplotype structure and linking correlated alleles of possible functional variants,” Bioinformatics (Oxford, England), vol. 31, no. 21, pp. 3555–3557, Dec. 2015, doi: 10.1093/BIOINFORMATICS/BTV402.

[54] K. Weissenborn, M. Bokemeyer, J. Krause, J. Ennen, and B. Ahl, “Neurological and neuropsychiatric syndromes associated with liver disease,” AIDS, vol. 19 Suppl 3, pp. S93–8, Oct. 2005.

[55] B. Sureka, K. Bansal, Y. Patidar, S. Rajesh, A. Mukund, and A. Arora, “Neurologic Manifestations of Chronic Liver Disease and Liver Cirrhosis,” Curr. Probl. Diagn. Radiol., vol. 44, no. 5, pp. 449–461, Sep. 2015.

[56] M. Pinter, M. Trauner, M. Peck-Radosavljevic, and W. Sieghart, “Cancer and liver cirrhosis: implications on prognosis and management,” ESMO Open, vol. 1, no. 2, p. e000042, Mar. 2016.

[57] “Mapping the human genetic architecture of COVID-19,” Nature 2021, pp. 1–8, Jul. 2021, doi: 10.1038/s41586-021-03767-x.

[58] D. A. Vyas, L. G. Eisenstein, and D. S. Jones, “Hidden in Plain Sight — Reconsidering the Use of Race Correction in Clinical Algorithms,” https://doi.org/10.1056/NEJMms2004740, vol. 383, no. 9, pp. 874–882, Jun. 2020, doi: 10.1056/NEJMMS2004740.

[59] N. D. Eneanya, W. Yang, and P. P. Reese, “Reconsidering the Consequences of Using Race to Estimate Kidney Function,” JAMA, vol. 322, no. 2, pp. 113–114, Jul. 2019, doi: 10.1001/JAMA.2019.5774.

[60] P. B. Fontanarosa and H. Bauchner, “Race, Ancestry, and Medical Research,” JAMA, vol. 320, no. 15, pp. 1539–1540, Oct. 2018, doi: 10.1001/JAMA.2018.14438.

[61] R. H. Kowalsky, A. C. Rondini, and S. L. Platt, “The Case for Removing Race From the American Academy of Pediatrics Clinical Practice Guideline for Urinary Tract Infection in Infants and Young Children With Fever,” JAMA Pediatrics, vol. 174, no. 3, pp. 229–230, Mar. 2020, doi: 10.1001/JAMAPEDIATRICS.2019.5242.

[62] Z. Obermeyer, B. Powers, C. Vogeli, and S. Mullainathan, “Dissecting racial bias in an algorithm used to manage the health of populations,” Science, vol. 366, no. 6464, pp. 447–453, Oct. 2019, doi: 10.1126/SCIENCE.AAX2342.

[63] N. Risch, E. Burchard, E. Ziv, and H. Tang, “Categorization of humans in biomedical research: genes, race and disease,” Genome Biology 2002 3:7, vol. 3, no. 7, pp. 1–12, Jul. 2002, doi: 10.1186/GB-2002-3-7-COMMENT2007.

[64] S. A. Tishkoff and K. K. Kidd, “Implications of biogeography of human populations for ‘race’ and medicine,” Nature Genetics 2004 36:11, vol. 36, no. 11, pp. S21–S27, Oct. 2004, doi: 10.1038/ng1438.

[65] D. Reich et al., “Reduced Neutrophil Count in People of African Descent Is Due To a Regulatory Variant in the Duffy Antigen Receptor for Chemokines Gene,” PLoS Genetics, vol. 5, no. 1, Jan. 2009, doi: 10.1371/JOURNAL.PGEN.1000360.

[66] S. A. Atallah-Yunes, A. Ready, and P. E. Newburger, “Benign Ethnic Neutropenia,” Blood reviews, vol. 37, Sep. 2019, doi: 10.1016/J.BLRE.2019.06.003.

[67] S. L. van Driest et al., “Association Between a Common, Benign Genotype and Unnecessary Bone Marrow Biopsies Among African American Patients,” JAMA Internal Medicine, vol. 181, no. 8, pp. 1100–1105, Aug. 2021, doi: 10.1001/JAMAINTERNMED.2021.3108.

[68] P. S. Rao et al., “A comprehensive risk quantification score for deceased donor kidneys: The kidney donor risk index,” Transplantation, vol. 88, no. 2, pp. 231–236, Jul. 2009, doi: 10.1097/TP.0B013E3181AC620B.

[69] B. A. Julian et al., “Effect of Replacing Race With Apolipoprotein L1 Genotype in Calculation of Kidney Donor Risk Index,” American Journal of Transplantation, vol. 17, no. 6, pp. 1540–1548, Jun. 2017, doi: 10.1111/AJT.14113.

[70] S. Limou, G. W. Nelson, J. B. Kopp, and C. A. Winkler, “APOL1 Kidney Risk Alleles: Population Genetics and Disease Associations,” Advances in chronic kidney disease, vol. 21, no. 5, p. 426, 2014, doi: 10.1053/J.ACKD.2014.06.005.

[71] N. Mars et al., “The role of polygenic risk and susceptibility genes in breast cancer over the course of life,” Nature Communications 2020 11:1, vol. 11, no. 1, pp. 1–9, Dec. 2020, doi: 10.1038/s41467-020-19966-5.

[72] M. G. Levin and D. J. Rader, “Polygenic Risk Scores and Coronary Artery Disease,” Circulation, vol. 141, pp. 637–640, Feb. 2020, doi: 10.1161/CIRCULATIONAHA.119.044770.

[73] M. J. Landrum et al., “ClinVar: Improving access to variant interpretations and supporting evidence,” Nucleic Acids Research, vol. 46, no. D1, pp. D1062–D1067, Jan. 2018.

[74] C. C. Chang, C. C. Chow, S. Tellier Laurent Cam and Vattikuti, S. M. Purcell, and J. J. Lee, “Second-generation PLINK: rising to the challenge of larger and richer datasets,” Gigascience, vol. 4, p. 7, Feb. 2015.

[75] A. Manichaikul, J. C. Mychaleckyj, S. S. Rich, K. Daly, M. Sale, and W.-M. Chen, “Robust relationship inference in genome-wide association studies,” Bioinformatics, vol. 26, no. 22, pp. 2867–2873, Nov. 2010.

[76] S. Das et al., “Next-generation genotype imputation service and methods,” Nat. Genet., vol. 48, no. 10, pp. 1284–1287, Oct. 2016.

[77] P.-R. Loh et al., “Reference-based phasing using the Haplotype Reference Consortium panel,” Nat. Genet., vol. 48, no. 11, pp. 1443–1448, Nov. 2016.

[78] D. Taliun et al., “Sequencing of 53,831 diverse genomes from the NHLBI TOPMed Program,” Nature, vol. 590, no. 7845, pp. 290–299, Feb. 2021.

[79] C. Fuchsberger, G. R. Abecasis, and D. A. Hinds, “minimac2: faster genotype imputation,” Bioinformatics, vol. 31, no. 5, pp. 782–784, Mar. 2015.

[80] G. Abraham, Y. Qiu, and M. Inouye, “FlashPCA2: principal component analysis of Biobank-scale genotype datasets,” Bioinformatics, vol. 33, no. 17, pp. 2776–2778, Sep. 2017.

[81] O. Delaneau, J.-F. Zagury, M. R. Robinson, J. L. Marchini, and E. T. Dermitzakis, “Accurate, scalable and integrative haplotype estimation,” Nature Communications 2019 10:1, vol. 10, no. 1, pp. 1–10, Nov. 2019, doi: 10.1038/s41467-019-13225-y.

[82] R. Shemirani, G. M. Belbin, C. L. Avery, E. E. Kenny, C. R. Gignoux, and J. L. Ambite, “Rapid detection of identity-by-descent tracts for mega-scale datasets,” Nature Communications 2021 12:1, vol. 12, no. 1, pp. 1–13, Jun. 2021, doi: 10.1038/s41467-021-22910-w.

[83] J. C. Denny et al., “Systematic comparison of phenome-wide association study of electronic medical record data and genome-wide association study data,” Nat. Biotechnol., vol. 31, no. 12, pp. 1102–1110, Dec. 2013.

