## Supplemental Materials for "Leveraging genomic diversity for discovery in an EHR-linked biobank: the UCLA ATLAS Community Health Initiative"

### Supplementary Material

| SIRE race | Race | Percentage (%) | N |
|---|---|---|---|
| African American | Black, Black or African American, Black: African American | 4.7 | 94,543 |
| Asian | Asian, Asian: Pakistani, Asian: Thai, Asian: Indonesian, Asian: Taiwanese, Vietnamese, Japanese, Asian Other, Filipino, Korean, Asian Indian, Asian: Vietnamese, Chinese, Asian: Japanese, Asian: Korean, Asian: Asian Indian, Asian: Filipino, Asian: Other, Asian: Chinese | 7.4 | 147,517 |
| White or Caucasian | White or Caucasian | 44.5 | 887,766 |
| Other Race | Other Race | 13.1 | 261,739 |
| American Indian | American Indian or Alaska Native, American Indian, Alaska Native | 0.4 | 8,035 |
| Pacific Islander | Guamanian or Chamorro, Hawaiian Native, Native Hawaiian or Other Pacific Islander, Pacific Islander Other, Pacific Islander: Guamanian or Chamor gro, Pacific Islander: Native Hawaiian, Pacific Islander: Other, Pacific Islander: Samoan, Samoan | 0.2 | 3,708 |
| Unknown race | 'Declined to Specify', 'Unknown', None, '*Unspecified' | 29.6 | 591,082 |

**Supplementary Table S1: Self-reported race groupings**

We group together multiple race categories when describing self-reported race/ethnicity (SIRE) for the UCLA Health patient population (N=1.9 million). The broader group names are noted in the first column and the list of specific races within each category are in the second column. Note that the percentages reported in the main text are computed without the inclusion of individuals with ‘Unknown race’.

| SIRE ethnicity | Ethnicity | Percentage (%) | N |
|---|---|---|---|
| Hispanic/Latino | Hispanic or Latino; Mexican, Mexican American, Chicano/a; Hispanic/Spanish origin Other; Puerto Rican, Cuban | 12.9 | 256,315 |
| Not Hispanic/Latino | Not Hispanic or Latino | 56.3 | 1,122,410 |
| Unknown ethnicity | Unknown, Patient Refused, Unspecified | 30.9 | 615,665 |

**Supplementary Table S2: Self-reported ethnicity groupings**

We group together multiple ethnicity categories when describing self-reported race/ethnicity (SIRE) for the UCLA Health patient population (N=1.9 million). The broader group names are noted in the first column and the list of specific ethnicities within each category are in the second column. Note that the percentages reported in the main text are computed without the inclusion of individuals with ‘Unknown ethnicity.

| SIRE | Abbreviation | Percentage (%) | N |
|---|---|---|---|
| Not Hispanic/Latino - White/Caucasian | NH-WC | 36.81 | 734,093 |
| Not Hispanic/Latino - Black/African American | NH-AfAm | 4.40 | 87,700 |
| Not Hispanic/Latino - Asian | NH-Asian | 6.88 | 137,153 |
| Not Hispanic/Latino - American Indian | NH-AmIn | 0.30 | 5938 |
| Not Hispanic/Latino - Pacific Islander | NH-PI | 0.15 | 3019 |
| Not Hispanic/Latino - Other Race | NH-Other | 6.31 | 125,882 |
| Not Hispanic/Latino - Unknown Race | NH-Unk | 1.44 | 28,625 |
| Hispanic/Latino - White/Caucasian | HL-WC | 5.60 | 111,699 |
| Hispanic/Latino - Black/African American | HL-AfAm | 0.13 | 2690 |
| Hispanic/Latino - Asian | HL-Asian | 0.14 | 2849 |
| Hispanic/Latino - American Indian | HL-AmIn | 0.09 | 1817 |
| Hispanic/Latino - Pacific Islander | HL-PI | 0.02 | 396 |
| Hispanic/Latino - Other Race | HL-Other | 5.87 | 117,094 |
| Hispanic/Latino - Unknown Race | HL-Unk | 0.99 | 19,770 |
| Unknown Ethnicity - White/Caucasian | Unk-WC | 2.10 | 41,974 |
| Unknown Ethnicity - Black/African American | Unk-AfAm | 0.21 | 4153 |
| Unknown Ethnicity - Asian | Unk-Asian | 0.38 | 7515 |
| Unknown Ethnicity - American Indian | Unk-AmIn | 0.01 | 280 |
| Unknown Ethnicity - Pacific Islander | Unk-PI | 0.02 | 293 |
| Unknown Ethnicity - Other Race | Unk-Other | 0.94 | 18,763 |
| Unknown Ethnicity - Unknown Race | Unk-Unk | 27.21 | 542,687 |

**Supplementary Table S3: SIRE overview at UCLA**

We construct self-reported race/ethnicity (SIRE) groupings by constructing pairs of all race and ethnicity combinations for all individuals that have at least 1 diagnosis code (N=1.9 million). See Supplementary Table 1 and Supplementary Table 2 for a further breakdown of race and ethnicity groupings used to construct SIREs. We report the abbreviation, percentage, and the sample size of each SIRE.

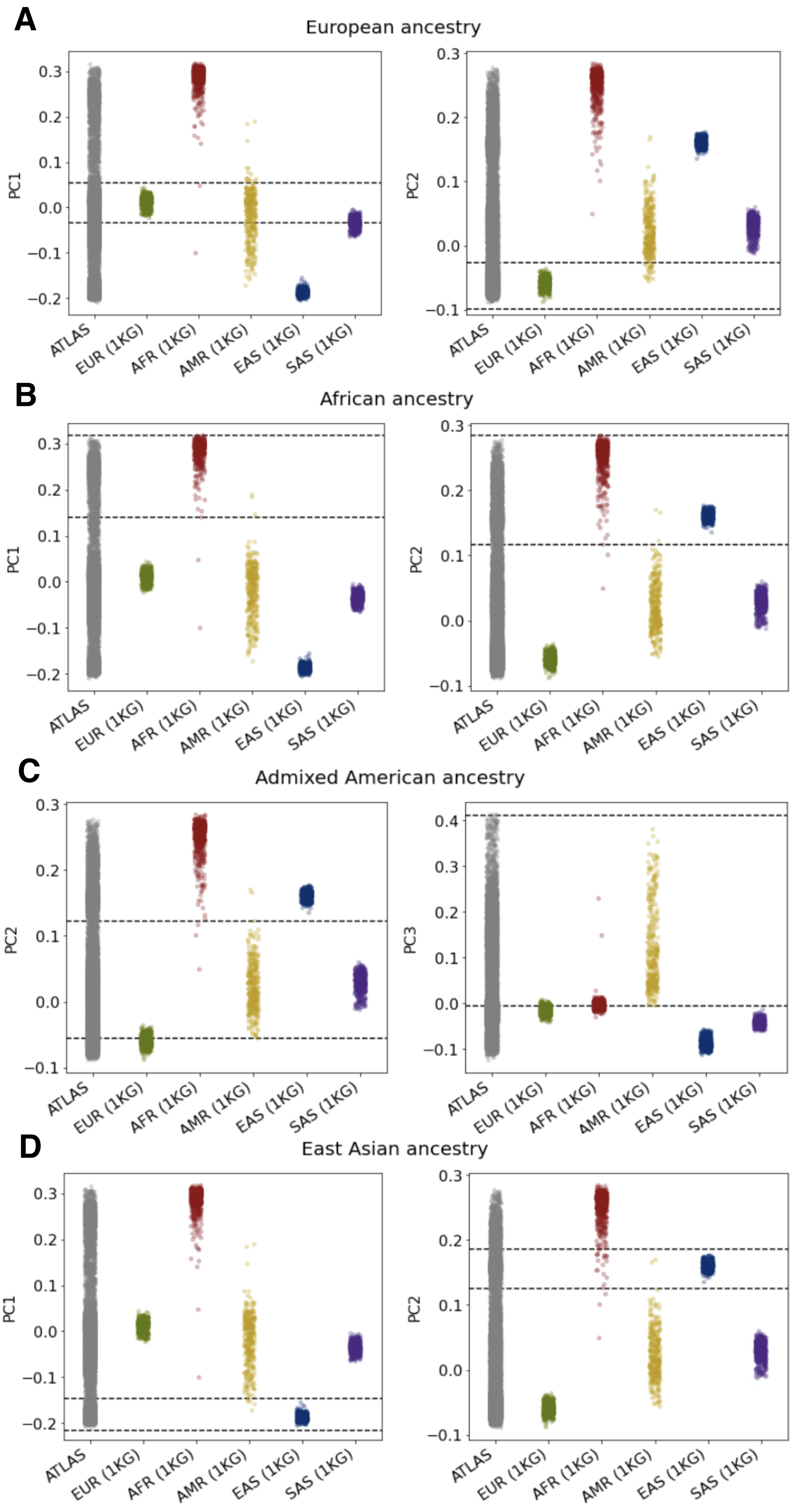
A
European ancestry
B
African ancestry
C
Admixed American ancestry
D
East Asian ancestry
PC1
PC2
PC3
ATLAS
EUR (1KG)
AFR (1KG)
AMR (1KG)
EAS (1KG)
SAS (1KG)

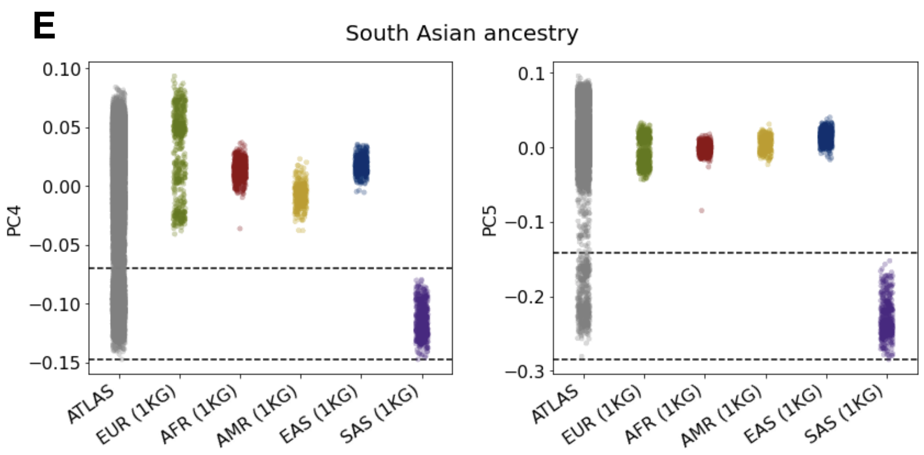


**Supplementary Figure S1: Clustering individuals by continental genetic ancestry using PCA-based clustering.** Genetic PCs of ATLAS participants (N=25,842) and individuals in 1000 Genomes shaded by continental ancestry: (A) European, (B) African, (C) Admixed American, (D) East Asian, (E) South Asian. Each continental ancestry cluster is described by two PCs. Dotted horizontal lines denote the threshold used to define each continental ancestry cluster.

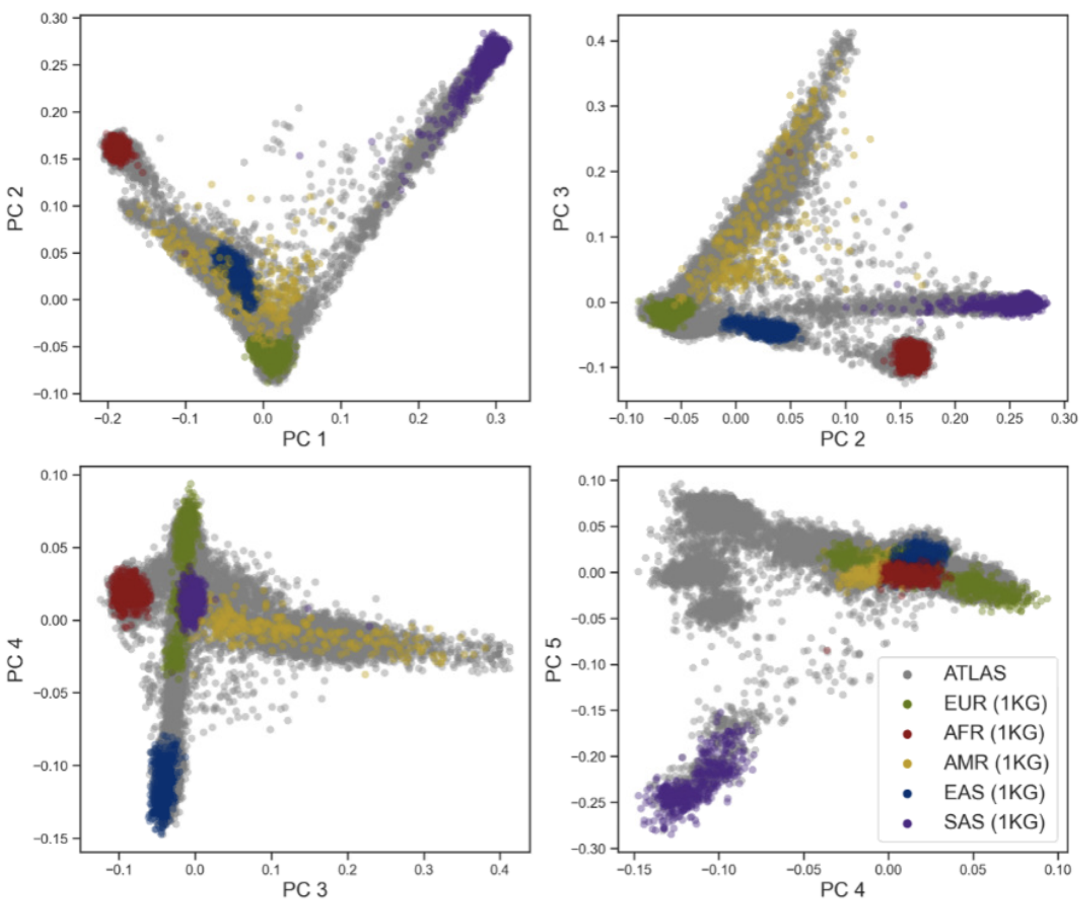


**Supplementary Figure S2: PCA-based clustering describes individuals by continental ancestry** Genetic PCs 1-5 of individuals in 1000 Genomes, colored by continental ancestry (European, African, Admixed American, East Asian, South Asian) and superimposed on the genetic PCs of the ATLAS participants (N=25,842).

| Continental ancestry | Abbreviation | Percentage (%) | N |
|---|---|---|---|
| European | EUR | 55.60 | 15,714 |
| African | AFR | 4.36 | 1,233 |
| Admixed Americas | AMR | 16.31 | 4,610 |
| East Asian | EAS | 8.36 | 2,362 |
| South Asian | SAS | 1.39 | 3,93 |
| Admixed or other ancestry | OTHER | 13.98 | 3,951 |

**Supplementary Table S4: Continental genetic ancestry in ATLAS.** We infer continental genetic ancestry for all unrelated individuals in ATLAS (N= 25,842) through principal component analysis (PCA) based clustering. Reference panels from 1000 Genomes was used for determining continental genetic ancestry. Individuals who were not able to be assigned to a single genetic ancestry category are denoted as ‘Admixed or other ancestry’.

| | EUR | AFR | AMR | EAS | SAS | N |
|---|---|---|---|---|---|---|
| NH-WC | 14519 | 13 | 657 | 19 | 19 | 16764 |
| NH-Afr | 6 | 1118 | 40 | 0 | 1 | 1426 |
| HL-Oth | 179 | 3 | 2059 | 3 | 0 | 2426 |
| HL-WC | 354 | 3 | 1464 | 1 | 0 | 1859 |
| NH-Asian | 21 | 1 | 4 | 2123 | 272 | 2901 |
| NH - American Indian | 38 | 2 | 28 | 0 | 1 | 67 |
| NH - Pacific Islander | 1 | 0 | 6 | 32 | 5 | 85 |

**Supplementary Table S5: Concordance between SIRE and continental genetic ancestry.**
Contingency table showing the sample overlap between self-reported race/ethnicity (SIRE) and inferred continental genetic ancestry of ATLAS individuals.

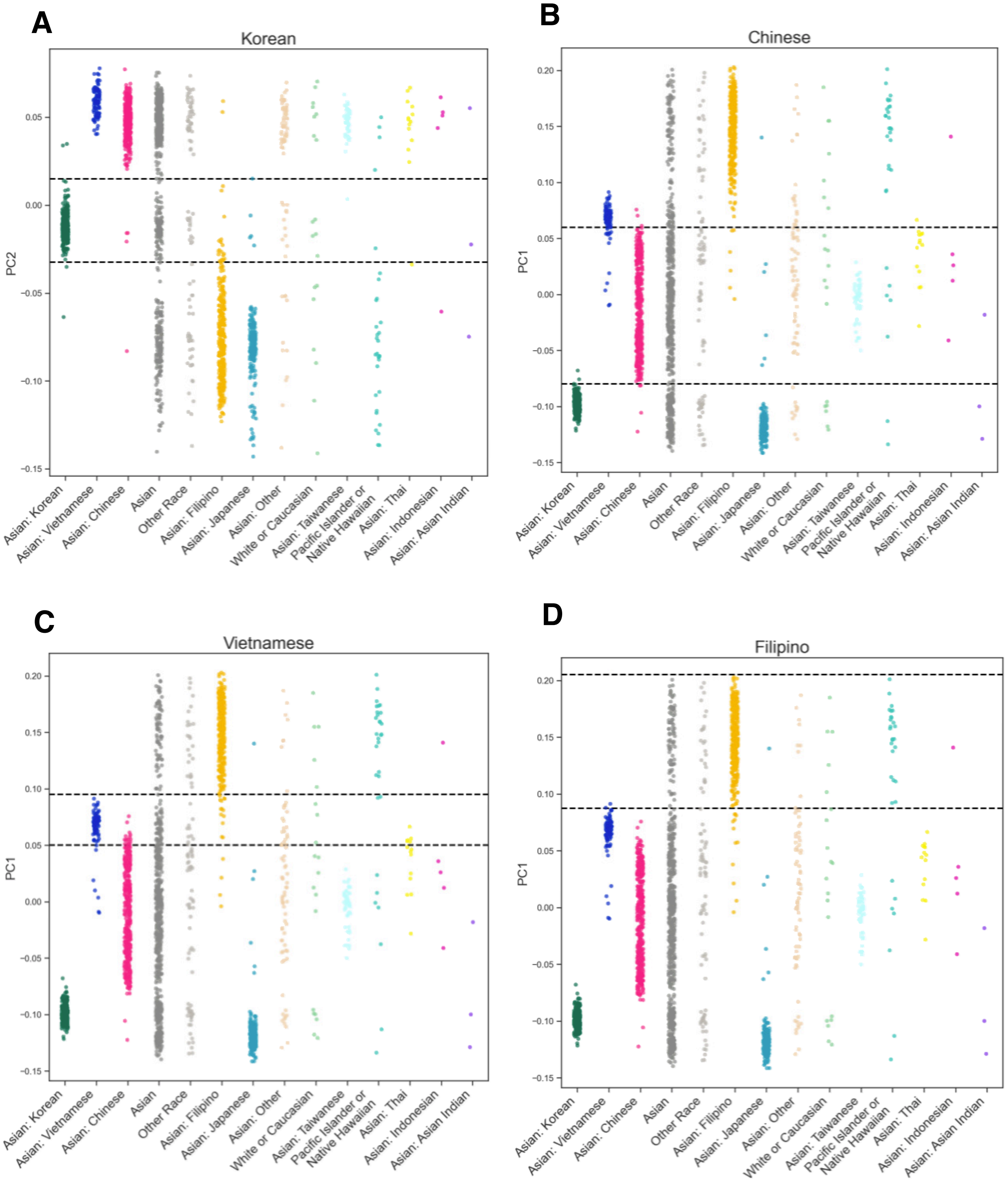

A
Korean
PC2
0.05
0.00
-0.05
-0.10
-0.15
B
Chinese
PC1
0.20
0.15
0.10
0.05
0.00
-0.05
-0.10
-0.15
C
Vietnamese
PC1
D
Filipino
PC1
Asian: Korean
Asian: Vietnamese
Asian: Chinese
Asian
Other Race
Asian: Filipino
Asian: Japanese
Asian: Other
White or Caucasian
Asian: Taiwanese
Pacific Islander or Native Hawaiian
Asian: Thai
Asian: Indonesian
Asian: Asian Indian

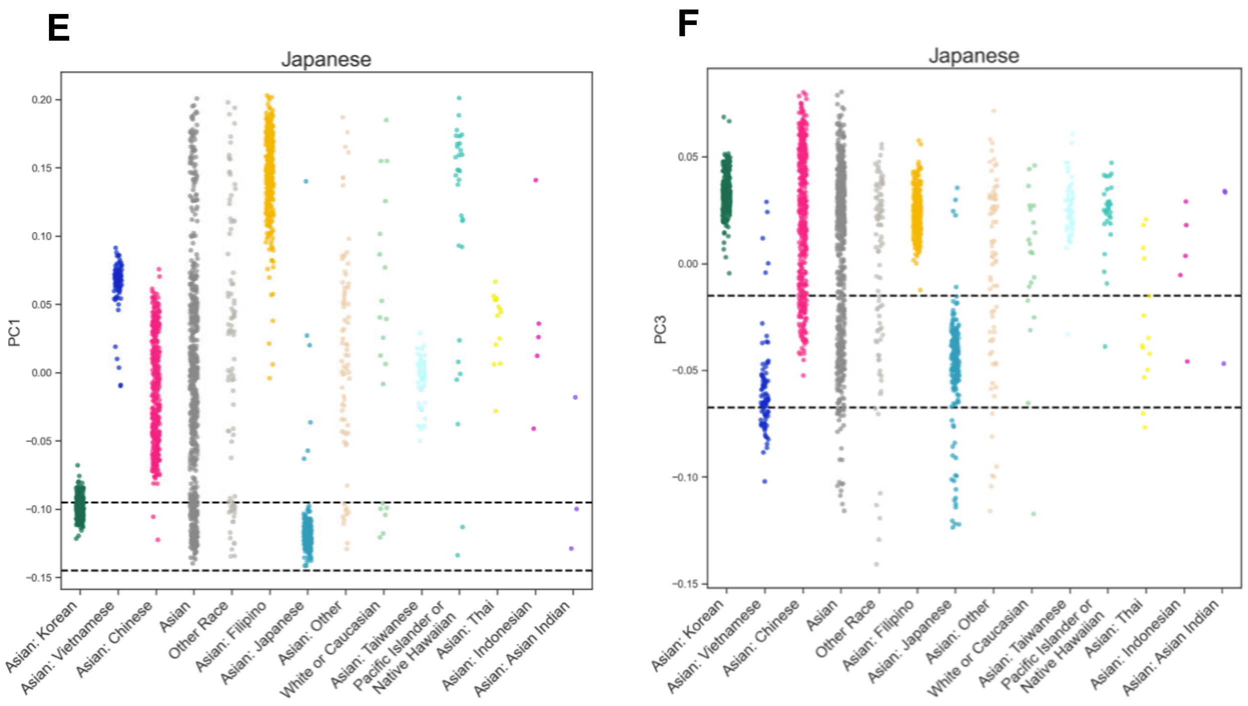


**Supplementary Figure S3: Clustering individuals by subcontinental genetic ancestry using PCA within the East Asian population.** Genetic PCs of East Asian ancestry ATLAS participants (N=2,242) colored by self-reported race: (A) Korean, (B) Chinese, (C) Vietnamese, (D) Filipino, (E, F) Japanese. Each subcontinental ancestry cluster is described by a single PC except for the Japanese population which uses two PCs (E, F). Dotted horizontal lines denote the threshold used to define each subcontinental ancestry cluster within ATLAS.

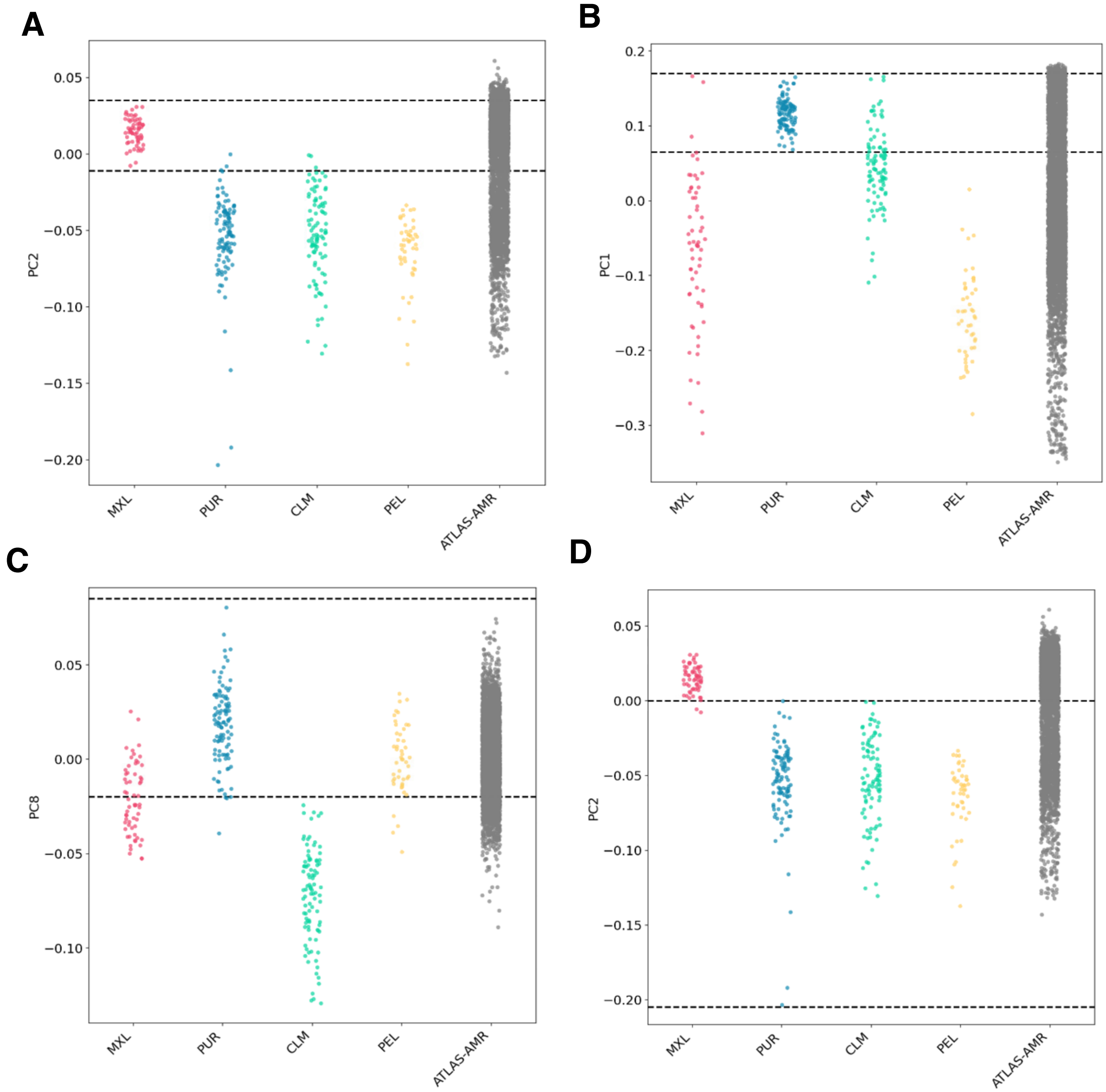


**Supplementary Figure S4: Clustering individuals by subcontinental genetic ancestry using PCA within the Admixed American population.** Genetic PCs of Admixed American ancestry ATLAS participants (N=4,597) and individuals of Mexican (MXL), Puerto Rican (PUR), Columbian (CLM), and Peruvian (PEL) ancestry from 1000 Genomes. The Mexican ancestry cluster in ATLAS is described by PC1 and the Puerto Rican cluster is determined by PCs 1, 2, and 8. Explicit thresholds could not confidently be drawn for the Columbian and Peruvian ancestry clusters in ATLAS.

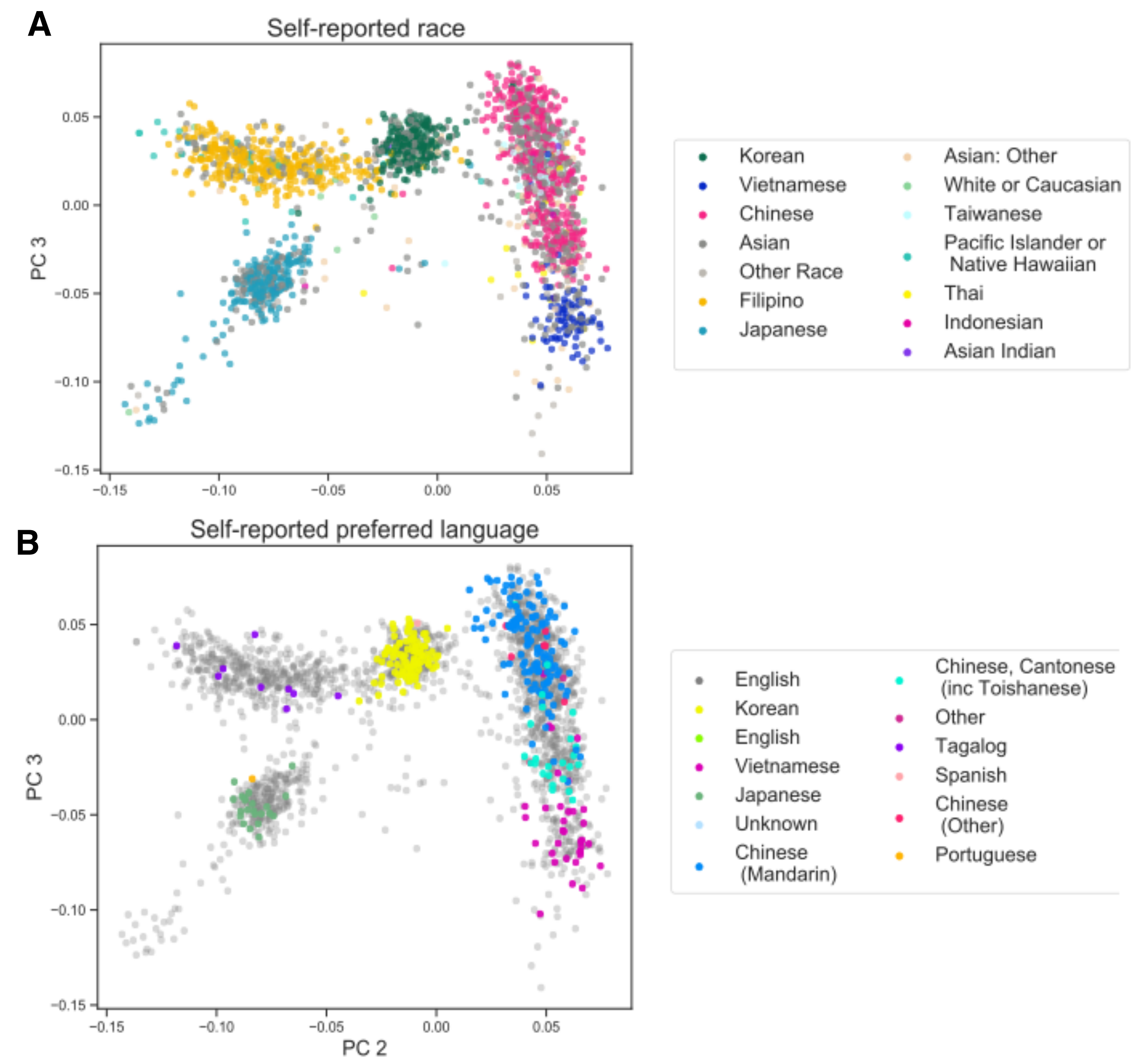


**Supplementary Figure S5: PCA of individuals with East Asian genetic ancestry in ATLAS by self-reported race and language.** (A) Genetic PCs 2 and 3 from principal component analysis performed on ATLAS-EAS individuals (N=2,242) colored by self-reported race and (B) self-reported preferred language. Only languages with >10 responses in ATLAS-EAS are assigned a color.

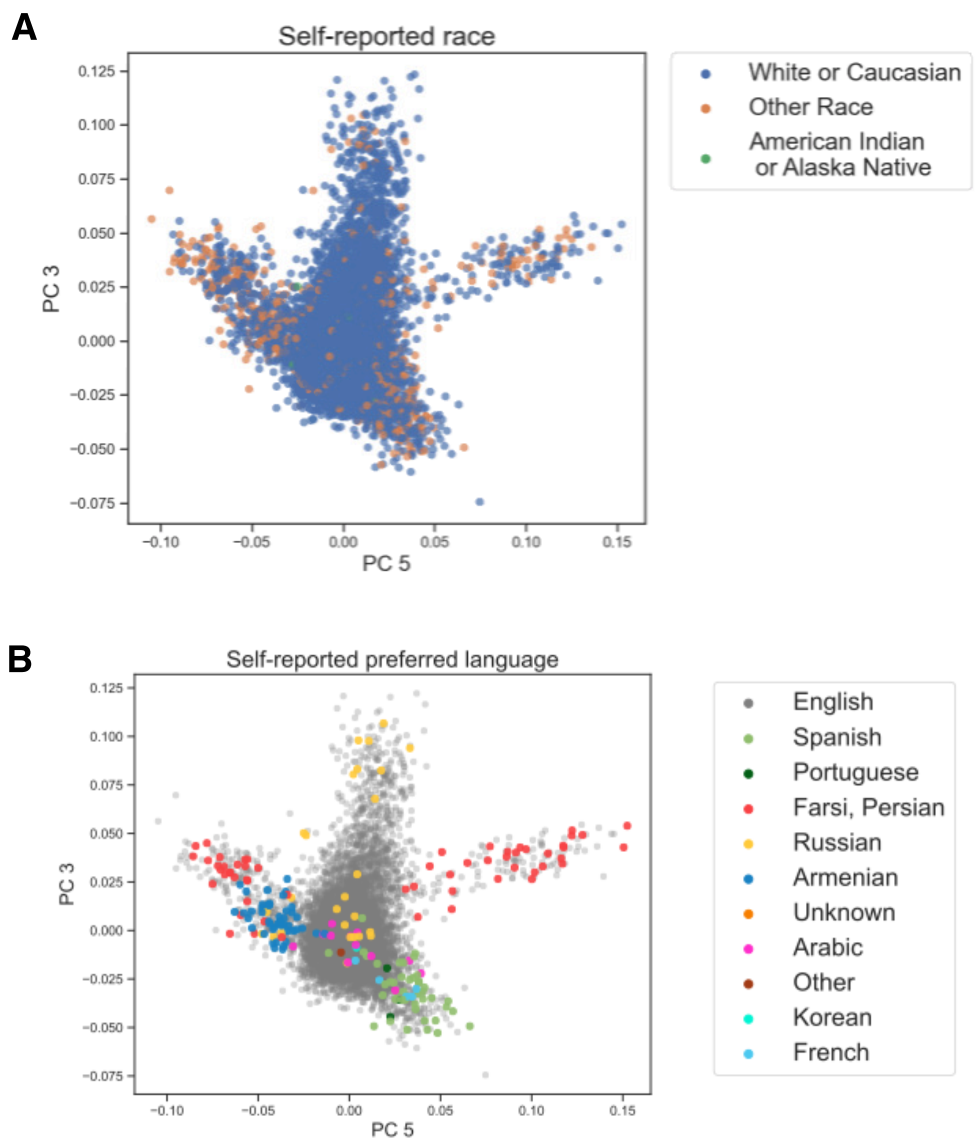


**Supplementary Figure S6: PCA of individuals with European genetic ancestry in ATLAS by self-reported race and language.** (A) Genetic PCs 3 and 5 from principal component analysis performed on ATLAS-EUR individuals (N=14,800). Only races and languages with >10 responses in ATLAS-EUR are assigned a color.

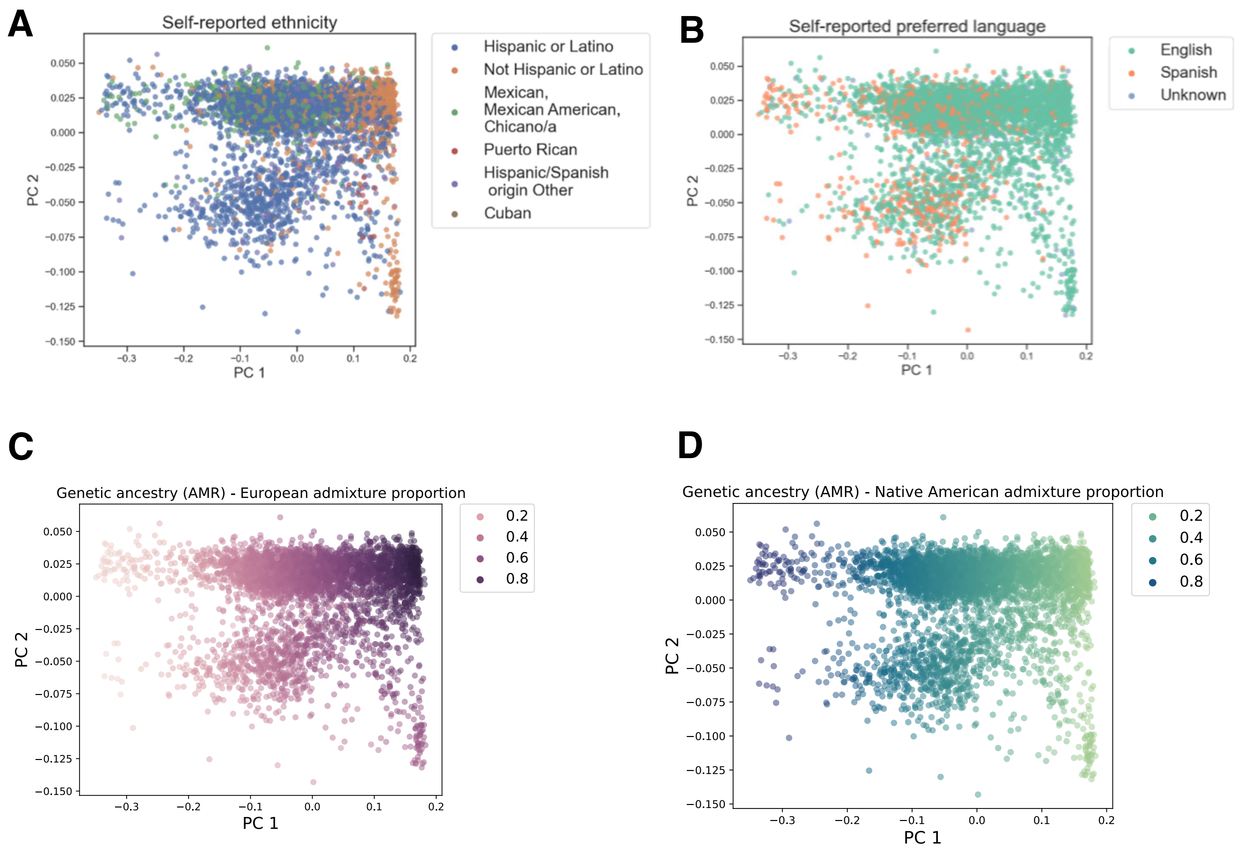


**Supplementary Figure S7: PCA of individuals with Admixed American genetic ancestry in ATLAS by self-reported ethnicity, preferred language, and estimated ancestry proportions.** (A) Principal component analysis performed on individuals in the Admixed American (AMR) continental ancestry group (N=4,597). (A) and (B) show PCs 1 and 2 for the ATLAS-AMR individuals colored by self-reported ethnicity and preferred language, respectively. Only languages with >10 responses in ATLAS-AMR are assigned a color. (C) and (D) show the PCs shaded according to the estimated proportion of European and Native American genetic ancestry inferred from ADMIXTURE.

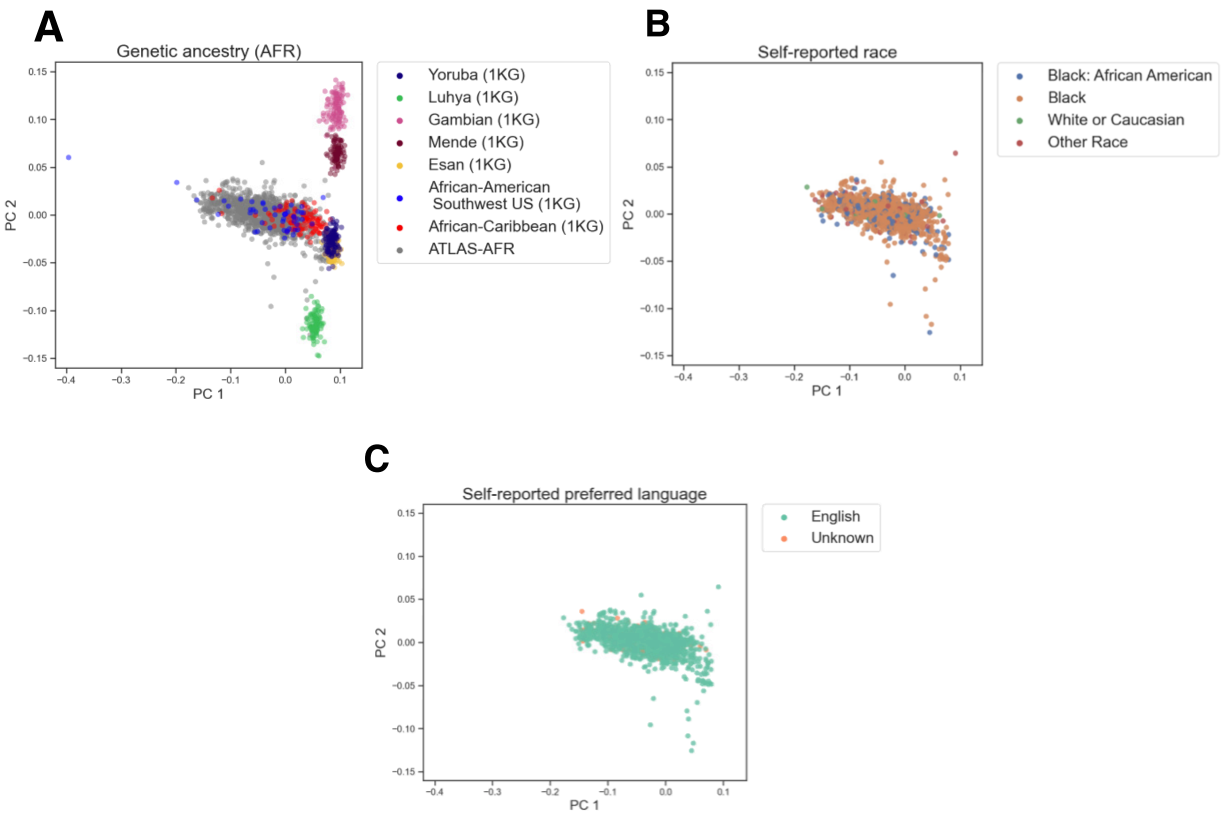


**Supplementary Figure S8: PCA of individuals with African genetic ancestry in ATLAS by subcontinental genetic ancestry, self-reported race, and preferred language.** (A) Principal component analysis performed on the African ancestry individuals from ATLAS (AFR-ATLAS) (N=1,257) and samples from the African subcontinental ancestry groups represented in 1000 Genomes. (A) Genetic PCs 1 and 2 where individuals from the 1000 Genomes African subcontinental ancestry groups are denoted by color and individuals from ATLAS are in gray. (B) and (C) show PCs 1 and 2 of ATLAS-AFR individuals colored by self-reported race and preferred language, respectively. Only languages with >10 responses in ATLAS-AFR are assigned a color.

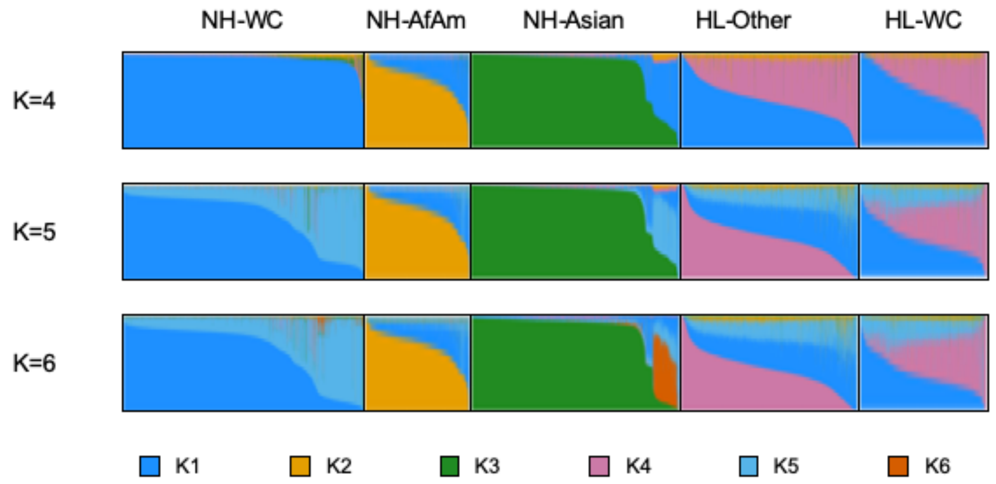


**Supplementary Figure 9: Individual admixture proportions vary among and within 5 self-reported race/ethnicity (SIRE) groupings.** Admixture proportions for ATLAS participants (N=25,872) were estimated using ADMXITURE with k=4, 5, or 6 ancestral populations. Within each SIRE grouping, individuals are ordered on the x-axis by global ancestry proportions.

| **SIRE** | **N** | **k1** | **k2** | **k3** | **k4** |
|---|---|---|---|---|---|
| NH-WC | 15,389 | 0.95 (0.08) | 0.01 (0.033) | 0.02 (0.055) | 0.02 (0.044) |
| NH-AfAm | 1313 | 0.24 (0.16) | 0.73 (0.17) | 0.01 (0.031) | 0.02 (0.023) |
| NH-Asian | 2665 | 0.12 (0.23) | 0.01 (0.032) | 0.85 (0.25) | 0.02 (0.018) |
| HL-Oth | 2206 | 0.49 (0.19) | 0.06 (0.054) | 0.01 (0.055) | 0.44 (0.2) |
| HL-WC | 1680 | 0.58 (0.22) | 0.05 (0.046) | 0.01 (0.034) | 0.36 (0.21) |
| NH-AmIn | 63 | 0.76 (0.27) | 0.05 (0.14) | 0.02 (0.034) | 0.17 (0.23) |
| NH-PI | 77 | 0.28 (0.26) | 0.03 (0.055) | 0.65 (0.3) | 0.04 (0.079) |

**Supplementary Table S7: Average ADMXITURE proportions stratified by SIRE.** Mean admixture proportions for ATLAS individuals (N= 25,872) stratified by SIRE. Standard deviations are reported in parentheses. The columns represent the sample sizes for each SIRE and the ancestry proportions: k1 (European), k2 (African), k3 (East Asian), k4 (Native American).

#### Supplementary Table S8: (see Excel sheet)

**Supplementary Table S8: Associations between continental genetic ancestry and EHR-derived phenotypes within ATLAS.** We perform an association test between the individuals' inferred genetic ancestry and 1,300 EHR-derived phenotypes (phecodes) across ATLAS (N= 25,872) while also adjusting for age and sex. Significance is determined after adjusting for the number of tested phenotypes ($p<3.8e-5$).

#### Supplementary Table S9: (see Excel sheet)

**Supplementary Table S9: Associations between genetic ancestry proportions and EHR-derived phenotypes within SIREs.** We perform a marginal regression between each of the ancestry proportions estimated from ADMIXTURE (k=4; European, African, East Asian, and Native American ancestry) and 1,300 EHR-derived phenotypes (phecodes) within each of the 7 ATLAS SIRE groups (NH-WC, NH-AfAm, HL-Other, HL-WC, NH-Asian, NH-PI, NH-AmIn). We additionally adjust for age and sex in the model. Only traits with >10 cases per SIRE were tested. Significance is determined after adjusting for the number of tested phenotypes ($p<3.8e-5$).

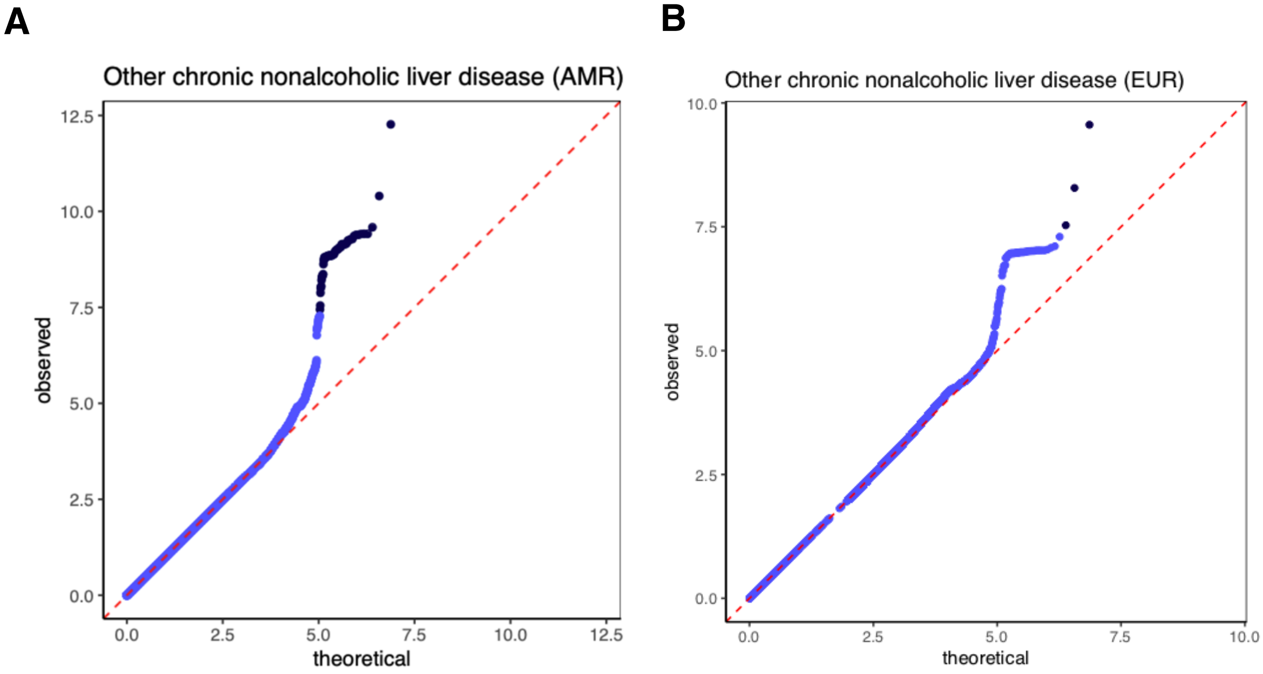


**Supplementary Figure S10: QQ-plots for Other chronic nonalcoholic liver disease GWAS in the AMR and EUR continental genetic ancestry group in ATLAS.** A) We performed a GWAS for 'Other chronic nonalcoholic liver disease' in the AMR continental ancestry group (N-Case: 919, N-Controls: 3262) using only common SNPs (MAF > 1%). The QQ-plot shows that the analysis is well-calibrated (lambda-GC: 1.01). Points in dark blue are SNPs that pass the genome-wide significance level (p<5e-08). B) We performed a GWAS for 'Other chronic nonalcoholic liver disease' in the EUR continental ancestry group (N-Case: 2,275, N-Controls: 14,155) using only common SNPs (MAF > 1%). The QQ-plot shows that the analysis is well-calibrated (lambda-GC: 1.02).

Supplementary Table S10: (see Excel sheet)

**Supplementary Table S10: Genome-wide significant associations for Other chronic nonalcoholic liver disease GWAS in the AMR and EUR continental genetic ancestry groups in ATLAS.** We performed a GWAS for 'Other chronic nonalcoholic liver disease' in the AMR continental ancestry group (N-Case: 919, N-Controls: 3,262) and the EUR continental ancestry group (N-Case: 2,275, N-Controls: 14,155) using only common SNPs (MAF > 1%). We provide the list of genome-wide significant associations (p<5e-8) for both populations.
